# The LAMp-LC Study: Unveiling the lung microbiome in Australians with early- and late-stage lung cancer using shotgun metagenomics

**DOI:** 10.64898/2026.11.10.26367577

**Authors:** Timothy Baird, Tiana R. Davey, Bryan A. Chan, Andrew Pattison, Olusola S. Olagoke, Miriana L. Littlejohn, K. Leigh Greathouse, Joshua K. Stone, Tamieka A. Fraser, Kasey A. Webb, Emily M. Dunn, Peter T. Bell, Andrew Dettrick, Michael Bint, Derek S. Sarovich, Erin P. Price

## Abstract

**Background:** Lung cancer remains the leading cause of cancerl71related mortality, yet the role of lower airway microbiota in disease development and progression is incompletely defined.

**Methods:** We characterised the lung microbiome in an Australian cohort (lung cancer *n*=106, healthy *n*=10) using shotgun metagenomics and semil71quantitative culture to evaluate microbial ecology across the tumour microenvironment (TME), contralateral airways, and oral cavity, to evaluate microbiome differences across earlyl71 and latel71stage disease, and to quantify clinical covariate impacts on airway microbiota. Diversity analyses, multivariable linear modelling, and culturel71based comparisons were performed, with adjustment for smoking packl71years, sex, and tumour characteristics.

**Results:** Cumulative smoking exposure caused significant, progressive *Neisseria subflava, Gemella sanguinis,* and *Rothia mucilaginosa* depletion, alongside *Streptococcus mitis* enrichment, with *Mogibacterium diversum* uniquely depleted within the TME of smokers. β diversity in both early- and late-stage bronchial washes (BWs) significantly diverged from healthy controls, whilst α diversity was significantly reduced in early-stage vs. late-stage BWs. The earlyl71stage TME demonstrated significantly reduced microbial cultivability compared with healthy and other lung cancer BWs. A small number of taxa were uniquely enriched (*Sporothrix* and *Pseudomonas* species) or depleted (*Streptococcus parasanguinis*, *Streptococcus cristatus*, *R. mucilaginosa*) within the TME, and 13 taxa were significantly enriched or depleted in both TME and contralateral airways compared with healthy airways. Only adenocarcinoma showed distinct oral βl71diversity restructuring.

**Conclusions:** The lung cancer microbiome is characterised by early and progressive lower airway dysbiosis that is strongly shaped by tumour presence. Our findings point to potential new avenues for microbiomel71based diagnostic and therapeutic strategies.

## Introduction

Lung cancer is the leading cause of cancer related mortality globally, and 1.8 million people are diagnosed every year^1^. Despite recent treatment advances, lung cancer remains an insidious disease with poor prognosis, with five-year survival rates of 27% (68% for Stage I and 3% for Stage IV disease)^2,3^. Given its largely asymptomatic nature until late-stage disease, coupled with its high mortality rate and significant socioeconomic burden, earlier diagnosis, improved therapeutic interventions, and prevention are essential for improving outcomes.

There is increasing recognition of the role microbes play in oncogenesis, with current estimates suggesting single-pathogen infections are linked to 12-16% of all human cancers^4,5^. However, major gaps remain in understanding the impact of microbial community changes (particularly microbiome dysbiosis) in lung cancer development, treatment response, and progression. Most lung cancer microbiome studies have used 16S rRNA variable-region amplicon sequencing (metataxonomics) to profile lung tissue and bronchoalveolar lavage fluid (BALF), and to a lesser extent, sputum^6^. Whilst these studies have reported the extratumoural or intratumoural enrichment of taxa such as *Veillonella*, *Streptococcus*, and *Acidovorax*^7–11^, enormous variability in study design, participant inclusion/exclusion criteria, specimen type/s, geographic region, and disease stage has made establishment of a consensus link challenging. A recent meta-analysis of intratumoural 16S metataxonomic studies attempted to overcome this variability, identifying broad shifts such as increased *Actinobacteria* and *Corynebacterium* in tumour tissues^12^. Nevertheless, 16S rRNA metataxonomics remains inherently limited by poor species-level resolution, the exclusion of viruses and fungi, and high susceptibility to contaminant amplification in low-biomass environments.

Recent investigations have shifted towards shotgun metagenomics to more comprehensively profile lung cancer microbiota in tumour tissue^13,14^ or BALF^15–19^. However, like 16S sequencing, metagenomic profiling has revealed a high degree of inter-study variability, with inconsistent alpha (α) and beta (β) diversity findings. Furthermore, no single microbe has been found to be enriched or depleted in lung cancer airways (tumour tissues or BALF specimens) compared with control cohorts. Like 16S-based studies, the lack of consensus is unsurprising due to differences in control groups (healthy individuals, people with benign lung diseases, normal adjacent lung tissues), statistical methodologies, clinical covariates, specimen collection and processing workflows, bioinformatic approaches, handling of background contamination, and environmental/geographical factors, along with small sample sizes. To address some of these challenges, large-scale multicentre analyses have attempted to map tumour-specific microbiomes across various cancers, including lung cancer^20,21^. One such study, by Nejman and colleagues, undertook 16S-based analysis of 1526 tumours across seven cancer types. Their analysis identified a significant enrichment of tobacco-degradation pathways in the lung tumours of smokers; however, due to the diverse and heterogeneous group of bacteria sharing these smoke-degrading metabolic capabilities, a uniform species-level signature was absent^20^. Concurrently, the recent retraction of a large, retrospective cancer microbiome study due to bioinformatic artefacts^22^ underscores the challenges of low-biomass sequencing and highlights the critical need for rigorous negative controls to account for kitome, bronchoscope, and upper respiratory tract contamination.

Although prior findings highlight the clinical potential of lung cancer microbiomics, significant knowledge gaps remain. Specifically, the field has struggled to untangle the confounding effects of cumulative tobacco exposure from tumour-driven dysbiosis, often failing to isolate a true disease signature from the background smoking microbiome. Furthermore, longitudinal mapping across early- and late-stage disease, particularly distinguishing localised changes within the tumour microenvironment (TME) from broader, systemic airway shifts, remains poorly defined. Reliance on sequencing alone without functional or culture-based validation has also obscured true biomass and viability fluctuations. Finally, no comprehensive shotgun metagenomic studies have been performed in Oceania lung cancer cohorts, hampering international generalisability due to unexplored geographical and cohort-specific variations.

To address these knowledge gaps, we employed shotgun metagenomics alongside semi-quantitative conventional culture to comprehensively characterise the extratumoural lung cancer microbiome in a prospective, single-centre cohort in Queensland (QLD), Australia. Our study had four primary aims: i) establish robust, contamination-controlled methods for metagenomic analysis of aseptically and prospectively collected bronchial washes (BWs); ii) define the ecology of oral and lung microbiota in people with lung cancer by comparing matched primary tumour BWs (i.e. the TME), contralateral healthy lung BWs, and oral washes with healthy controls; iii) evaluate microbiome shifts across early-stage (Stage I/II) and late-stage (Stage III/IV) disease using metagenomics and conventional culture; and iv) use rigorous multivariable modelling to statistically decouple the background smoking microbiome from true cancer-associated dysbiosis, while concurrently evaluating the impacts of sex, age, and histological subtype.

## Methods

### Study participants

One-hundred and ten individuals ≥18 years undergoing routine bronchoscopy for suspected lung cancer at Sunshine Coast University Hospital (Birtinya, QLD, Australia) between June 2020 and December 2025 were prospectively recruited. Individuals who previously or were currently receiving chemotherapy, radiotherapy, and/or immunotherapy, had undergone prior surgery or biopsy for lung cancer, or had received systemic antibiotics <3 months prior were excluded. A further ten subjects (9 with no known lung disease and 1 with a confirmed benign nodule) undergoing routine bronchoscopy were recruited as healthy controls (**Figure 1**). Six additional healthy controls provided oral washes only. Four suspected lung cancer subjects subsequently diagnosed with non-malignancy, and one subject with benign lung disease, were excluded from the study, leaving 106 in the evaluable cohort. Tumour staging was undertaken as described in Supplemental Methods.

**Figure 1.**
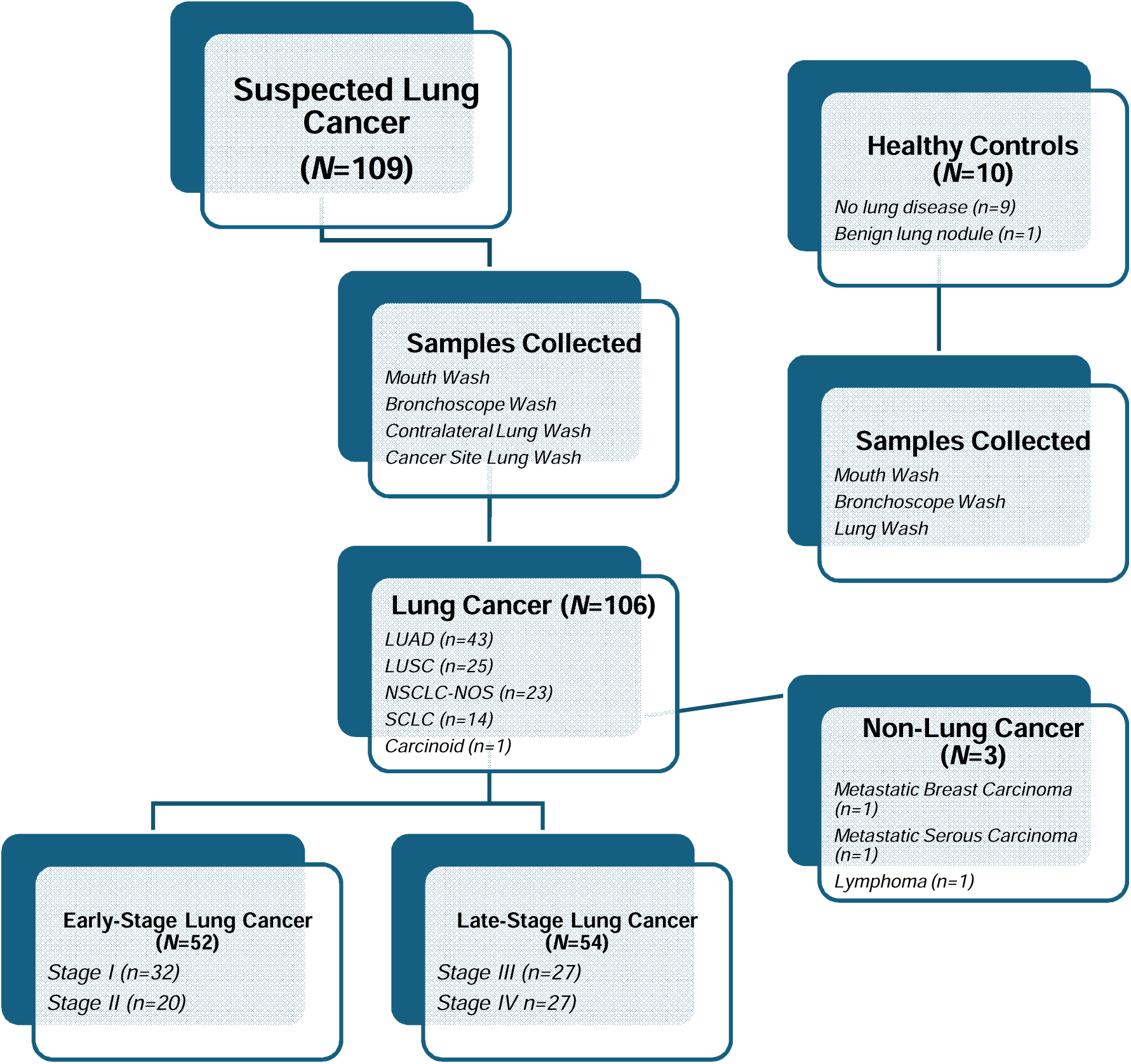
LAMp-LC Study flow chart outlining subject recruitment and sample collection. Abbreviations: LUAD, lung adenocarcinoma; LUSC, lung squamous cell carcinoma; NSCLC-NOS, non-small cell lung carcinoma, not otherwise specified; SCLC, small cell lung carcinoma. Not all enrolled participants underwent shotgun metagenomics; specimens failing DNA yield thresholds or quality controls were excluded (see Tables S1-S3 for the complete breakdown of sequenced specimens across mucosal sites, and negative controls).

### Specimen collection

Specimens were collected into sterile specimen pots in the following order during bronchoscopy: i) oral wash (∼10mL sterile phosphate-buffered saline [PBS]), collected immediately prior to sedation, to define the oral microbiome; ii) pre-procedure bronchoscope wash (∼10 mL PBS), collected immediately prior to bronchoscopy (*n*=44), to act as reagent-only controls; iii) BW (∼10-40 mL PBS) from the contralateral lung site (i.e. non-cancer side for early-stage cases, or the non-primary tumour side for late-stage cases), to provide within-cohort lung microbiome controls; and iv) BW (∼10-40 mL PBS) from the TME to profile extratumoural microbiota (**Figure 1**). Between the contralateral and TME BW collection, the bronchoscope was flushed with 10 mL PBS to minimise cross-contamination between lung sites. All specimens were collected prior to tumour tissue biopsy to mitigate the risk of intratumoural microbiome contamination. For the healthy control cohort, oral washes, scope washes, and BWs (from either the right or left upper lobe) were collected. All washings were stored at 4°C until processed. In total, 293 specimens were aseptically obtained from 106 confirmed lung cancer subjects: 91 oral washes, 101 contralateral BWs, and 101 TME BWs. A further 10 BWs and 15 oral washes were obtained from the healthy control cohort. Nine reagent-only controls and 24 bronchoscope washes were also subjected to metagenomic sequencing to account for kitome contamination.

### Specimen processing and microbiological culture

All specimens were pelleted via centrifugation at 4,667 x *g* for 10 minutes at 4°C; excess liquid was decanted into a fresh 3% Virkon solution, with ∼800 µL specimen resuspended via flick-mixing. Twenty µL was aliquoted directly onto MacConkey, Sabouraud dextrose, chocolate, Luria-Bertani, and mannitol salt agars, and subjected to the dilution streak technique out to a 5^th^ dilution streak. Culture plates were aerobically incubated at 37°C for up to 8 days to assess microbial growth.

#### Shotgun metagenomic sequencing and data analysis

DNA was obtained as detailed in Supplemental Methods. Illumina 150 bp paired-end libraries were sequenced on the NovaSeq 6000 (Australian Genome Research Facility, St Lucia, QLD, Australia) or NovaSeq X (AZENTA, Suzhou, China) platforms. Reads were filtered with Trimmomatic v0.39^23^ using previously described parameters^24^. The Kraken2 v2.17.1^25^ Standard v2/26/2026 database was used to flag ‘taxid 9606’ (i.e. *Homo sapiens*) and ‘taxid 9605’ (i.e. *Homo*) reads, which were then depleted using Seqtk v1.3 (https://github.com/lh3/seqtk). Additional contaminant taxa were also depleted prior to analysis (Supplemental Methods). Depleted metagenomic reads have been deposited under BioProject PRJNA1539961.

#### Microbial cultivability assessment

Chocolate agar is a non-selective, nutrient-rich, general-purpose medium that supports the growth of a broad range of respiratory microbes, including fastidious species like *Haemophilus influenzae*. This medium therefore provides a robust measure of the cultivable microbial load of aerobic and aerotolerant species in clinical specimens. Chocolate agar plates were prepared by four laboratorians using a standardised specimen volume (20 μL) to reduce inter-operator variability. Microbial growth was semi-quantitatively scored at Day 7-8 of incubation from 0 to 6, with 0 representing no growth, and 6 representing dense growth extending into the fifth dilution streak.

#### Statistical assessment

All sequence-based statistical analyses were undertaken using R (v4.6.0), executed in RStudio (v2026.06.0 build 242), and using R packages ggpubr^26^, ggrepel^27^, ggtext^28^, ggVennDiagram^29^, maaslin3^30^, patchwork^31^, phyloseq^32^, scales^33^, tidyverse^34^, and vegan^35^. Differences in multivariate dispersion were assessed using the PERMDISP2 procedure^36^ via the betadisper function within the vegan package. Paired datasets were: contralateral or TME BW vs. oral wash, and contralateral BW vs. TME BW (early-stage, late-stage). All other comparisons were unpaired. α and β diversity analyses, MaAsLin3^30^ mixed- and fixed-effects modelling, and culture-based statistical analyses, were undertaken as detailed in Supplemental Methods. We acknowledge the use of AI (Google Gemini 3 Series) for R code refinement.

### Ethics and safety

Human ethics and site-specific approvals were obtained from the Metro North Human Research Ethics Committee (ID: HREC/2021/QPCH/6950) and the Sunshine Coast Hospital and Health Service Research Governance Office. All participants provided written informed consent.

## Results

### Demographics

Mean ages for lung cancer (*n*=106; 71.9±8.1 years [**Table 1; Table S1**]) and control (*n*=10; 67.1±18.6 years [**Table S2**]) cohorts were similar; both were also gender-balanced (51.9% females vs. 50% males, respectively). Most lung cancer subjects (*n=*101; 95.3%) were current or former smokers (mean smoking pack-years=31.8±16.5 years). Similarly, most control subjects were former smokers (*n=*6; 60%) albeit with much lower smoking pack-years (8.3±2.6 years). Mean FEV_1_% was lower in the lung cancer cohort (77.6±19.4% vs. 90.2±6.4%). The most common primary tumour location was right upper lobe (*n*=39; 36.8%), followed by left upper lobe (*n*=29; 27.4%), left lower lobe (*n*=17; 16%) right lower lobe (*n*=15; 14.2%), and right middle lobe (*n*=6; 5.7%) **(Table 1)**.

**Table 1.** Summary of LAMp-LC Cohort with baseline demographic and clinical data.

| **Characteristic** | **Total (N=106)** |
|---|---|
| ***Age*** |  |
| Mean (±SD; Range) | 71.9 (8.1; 51-90) |
| ≤69 | 44 (41.5%) |
| ≥70 | 62 (58.5%) |
| ***Sex*** |  |
| Male | 55 (51.9%) |
| Female | 51 (48.1%) |
| ***Smoking Status*** |  |
| Never | 5 (4.7%) |
| Current/Ex-smoker | 101 (95.3%) |
| Pack-years (mean (±SD)) | 31.8 (16.51) |
| ***Lung Function (FEV<sub>1</sub>%)*** |  |
| Mean (±SD) | 77.6% (19.35) |
| ***Lung Cancer Site*** |  |
| Right upper lobe | 39 (36.8%) |
| Right middle lobe | 6 (5.7%) |
| Right lower lobe | 15 (14.2%) |
| Left upper lobe | 29 (27.4%) |
| Left lower lobe | 17 (16.0%) |
| ***Lung Cancer Subtype*** |  |
| Adenocarcinoma | 43 (40.5%) |
| Squamous cell carcinoma | 25 (23.6%) |
| NSCLC-NOS | 14 (13.2%) |
| Small cell carcinoma | 23 (21.7%) |
| Carcinoid | 1 (0.9%) |
| ***Lung Cancer Stage*** |  |
| Early (stages I-II) | 52 (49.1%) |
| Stage I | 32 (30.1%) |
| Stage II | 20 (18.9%) |
| Late (stages III-IV) | 54 (50.9%) |
| Stage III | *27* (25.5%) |
| Stage IV | *27* (25.5%) |
| ***Lung Cancer Gene Alteration*** |  |
| *EGFR* | 4 (3.8%) |
| *KRAS* | 15 (14.2%) |
| *BRAF* | 5 (4.7%) |
| *ROS1* | 1 (0.9%) |
| *PD-L1* negative (<1%) | 37 (34.9%) |
| *PD-L1* low (1-50%) | 20 (18.9%) |
| *PD-L1* high (>50%) | 17 (16.0%) |

Abbreviations: SD, one standard deviation; FEV1 (%), percentage of total air exhaled in the first second of a forced breath (FEV<sub>1</sub>) compared to total forced vital capacity (FVC); LUAD, lung adenocarcinoma; LUSC, lung squamous cell carcinoma; NSCLC-NOS, non-small cell lung carcinoma, not otherwise specified; SCLC, small cell lung carcinoma; *EGFR*, Epidermal Growth Factor Receptor gene; *KRAS*, Kirsten Rat Sarcoma virus oncogene homolog gene; *BRAF*, B-Raf proto-oncogene, serine/threonine kinase gene; *ROS1*, ROS proto-oncogene 1, receptor tyrosine kinase gene; *PD-L1*, Programmed Death-Ligand 1 gene.

### Lung cancer stages, subtypes and genotypes

Approximately equal numbers of participants with early-(49.1%: Stage I; *n*=32; Stage II; *n*=20) or late-(50.9%: Stage III; *n*=27; Stage IV; *n*=27) stage disease were recruited (**Figure 1**; **Table 1; Table S1**). Non-small cell lung carcinoma (NSCLC) was most common (*n*=91; 77.4%), followed by small-cell lung carcinoma (SCLC; *n*=14; 21.7%) and pulmonary carcinoid tumour (*n*=1; 0.9%). Histological subtyping identified lung adenocarcinoma (LUAD) (*n*=43; 40.5%) and lung squamous cell carcinoma (LUSC) (*n*=25; 23.6%) as the most common diagnoses following immunohistochemical staining, whereas 23 (17.1%) NSCLC cases were unable to be histologically classified/subtyped (NSCLC-NOS) (**Table 1**). Regarding genomic alterations identified, *KRAS* was most common (14/52; 26.9%), followed by *BRAF* (5/51; 9.8%), *EGFR* (4/54; 7.4%), *ROS1* (1/51; 2.0%), and *ALK* (1/53; 1.9%). *PD-L1* testing was negative (<1%), low (1-49%), and high (≥50%) in 37/74 (50.0%), 20/74 (27.0%), and 17/74 (23.0%) cases, respectively (**Table 1**).

### Cumulative tobacco smoking is associated with lung microbiome alterations

To determine the impacts of clinical confounders on lung microbiota, we included age, primary tumour location, biological sex, and tobacco exposure as variables across our BW dataset (**Table S1**). Using multivariable testing, age and primary tumour location did not significantly contribute to global microbial variance or individual taxon abundance/prevalence differences (FDR *q*>0.05) and were therefore excluded from subsequent models. Using a multivariable linear model adjusting for clinical cohort, sex, and tobacco exposure, cumulative smoking history (measured in pack-years) exerted the most profound influence on the lung microbiome. Longer pack-years history was associated with significant depletion in the prevalence of three core respiratory commensals: *Neisseria subflava* (β=-0.75), *Gemella sanguinis* (β=-0.72), and *Rothia mucilaginosa* (β=-0.67) (FDR *q*<0.05; **Figure 2)**. Whilst falling just outside the strict FDR boundary (*q*=0.055), *Streptococcus mitis* demonstrated a highly significant individual positive correlation with non-zero abundance based on cumulative smoking exposure (β=+1.05; *p*<0.001). Similarly, when examining biological sex, *Bradyrhizobium betae* demonstrated higher prevalence in male participants (β coefficient=+1.44) but fell just outside the strict FDR boundary (*q*=0.055). However, sex did not impact the smoking microbiome signature, with *N. subflava*, *G. sanguinis*, and *R. mucilaginosa* remaining significantly depleted.

**Figure 2.**
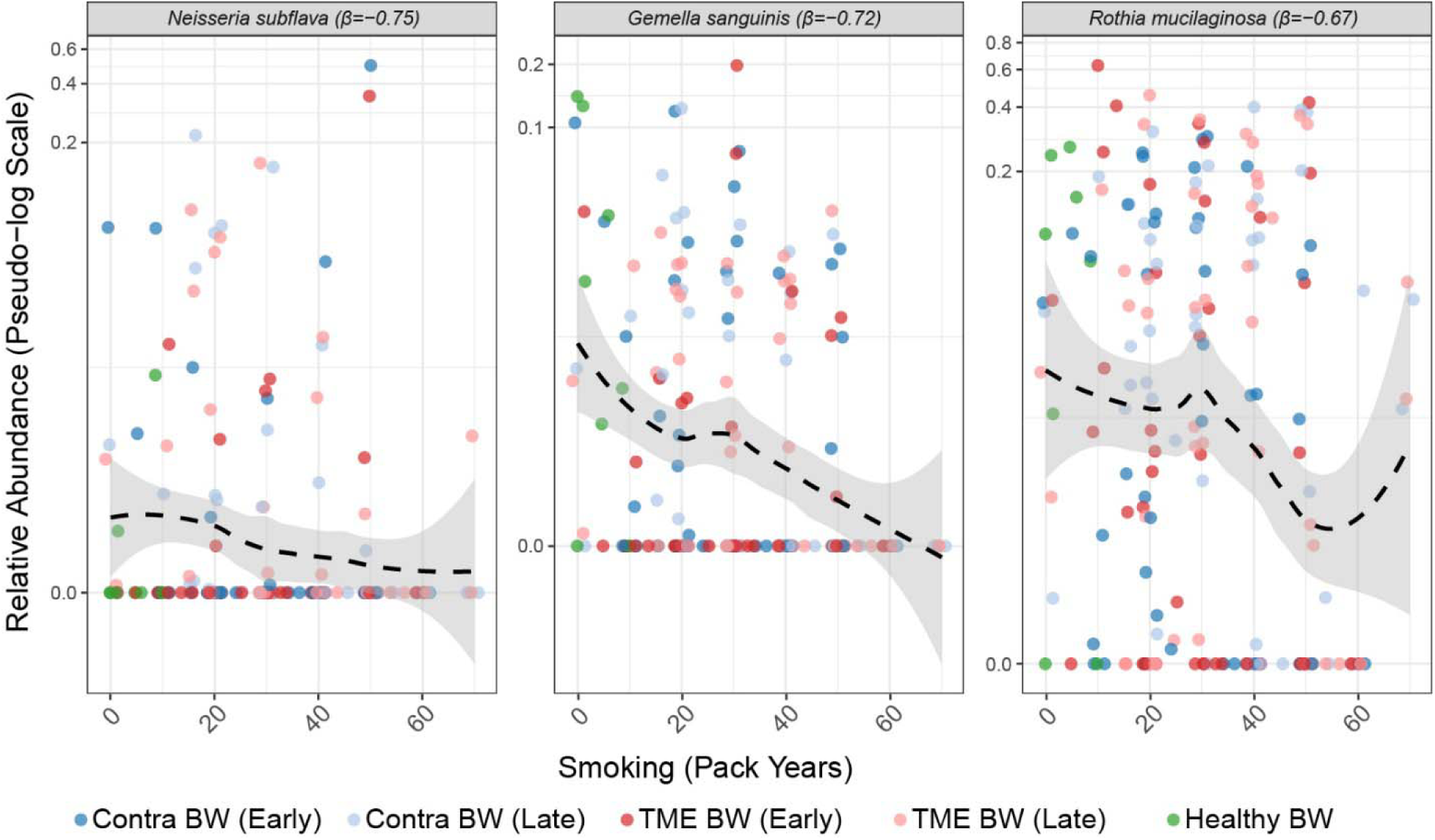
Impact of cumulative tobacco smoking on the core lung microbiome. Multivariable linear modelling (MaAsLin3) of the global clinical cohort (healthy control and lung cancer participants across all stages, sites, and histological subtypes) identified significant depletion of six core commensal taxa (β coefficients=−0.78 to −0.60) in heavier smokers. However, this model could not parse apart whether these significant taxa were associated with lung cancer due to a lack of healthy controls with long smoking pack year histories. Individual points represent single bronchial wash (BW) samples, coloured by clinical cohort (Contra=contralateral lung, TME=tumour microenvironment) and stage (Early=Stage I-II, Late=Stage III-IV). Dashed black lines indicate LOESS smoothing fits, with shaded grey areas representing the 95% confidence interval. Relative abundances are plotted on a pseudo-log scale (vertical axes). All depicted taxa exhibit an individual false discovery rate-adjusted *q*<0.05 after simultaneous statistical adjustment for the clinical cohort (fixed effect).

### Localised Mogibacterium diversum depletion requires a combination of cumulative tobacco exposure and tumour presence

We next investigated the presence of a tobacco smoking-associated microbiome strictly within our lung cancer cohort. By separately analysing TME BWs and contralateral BWs using fixed-effects modelling, we identified significantly lower *M. diversum* abundance in the TME of heavier smokers (β coefficient=-1.17; *q*<0.05; **Figure S1A**), but not in the paired contralateral lung (β coefficient=-0.40; *q*=0.98; **Figure S1A & B**). This smoking-driven depletion within the TME was robust whether independently controlling for tumour subtype (β coefficient=-1.17, FDR *q*=0.045) or disease stage (β coefficient=-1.05, FDR *q*=0.021). Interestingly, a paired mixed-effects analysis of the 88 matched BWs did not find a universally lower absolute abundance of *M. diversum* in the TME when compared directly to the contralateral airway (FDR *q*>0.05), confirming that tumour presence alone is not associated with *M. diversum* depletion. Taken together, these findings demonstrate that *M. diversum* depletion is associated with a combination of cumulative tobacco exposure and tumour presence.

### Global microbial community architecture shifts in lung cancer airways are associated with disease presence and disease stage, but not by anatomical site

Assessment of overarching microbial ecological differences between healthy and lung cancer cohorts using multivariable PERMANOVA (adjusted for pack-years) demonstrated significant global divergence in species composition (β diversity) between cohorts (*p*=0.022) (**Figure S2A**). Importantly, beta-dispersion testing confirmed equal variance between these cohorts (beta-dispersion *p*=0.678), confirming that this significance is driven by a genuine compositional shift rather than unequal ecological scatter. In contrast, unadjusted pairwise comparison of the Shannon diversity index identified similar α diversity between cohorts (*p*>0.05) (**Figure S2B**).

To ascertain whether this global disruption in lung cancer airways was associated with specific site-specific or clinical factors, we stratified our lung cancer cohort by anatomical site (contralateral or TME) or disease stage (early or late). Global disease stage analysis identified significant β diversity differences between healthy and stage-stratified lung cancer BWs (i.e. pooled contralateral and TME BWs; PERMANOVA *p*=0.004; **Figure 3A**). Notably, this global divergence was characterised not only by shifts in community composition, but by significant ecological destabilisation, with both early and late disease stages exhibiting significantly higher microbial variability than the tightly clustered Healthy baseline (beta-dispersion *p*=0.004). Importantly, multivariable modeling confirmed that this progressive structural disruption is driven by advancing disease stage, independent of cumulative smoking exposure. Subsequent post-hoc pairwise testing confirmed that the lower airway microbiome exists on a progressive continuum, with significant compositional shifts occurring not only between healthy controls and early-stage disease (FDR *p*=0.029) but continuing to diverge significantly as the disease advances to late-stage (Early vs. Late, FDR *p*=0.018). Furthermore, Kruskal-Wallis omnibus testing revealed significant α diversity differences between disease stages, confirmed with pairwise testing to be driven by a significantly lower Shannon diversity score in early-stage BWs compared with late-stage BWs (*p*=0.012; **Figure 3B**).

**Figure 3.**
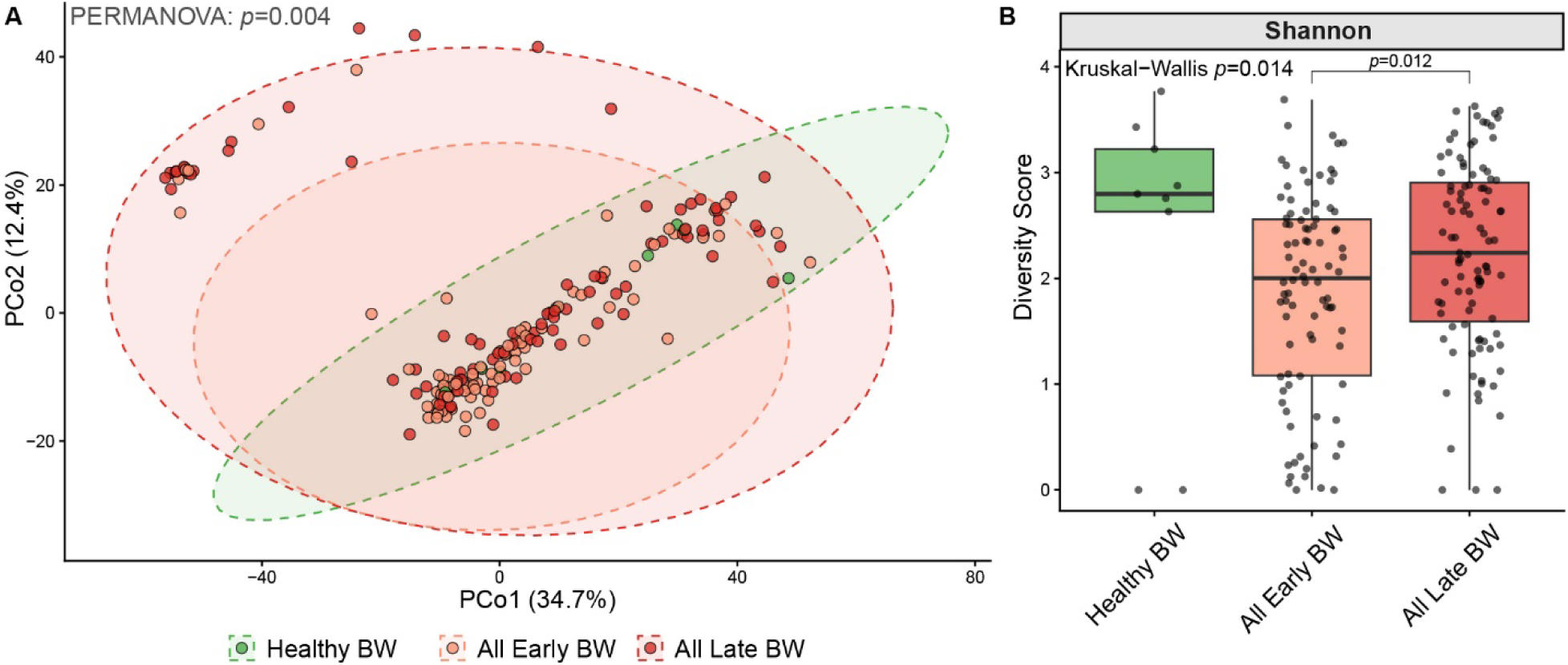
Disruption of lung microbiome diversity and structure is driven by disease stage. (A) Principal Coordinate Analysis based on Aitchison distance was used to visualise lung microbiome β diversity in healthy bronchial washes (BWs) and lung cancer BWs (i.e. contralateral and tumour microenvironment BWs) collected at early-or late-stage disease. Ellipses represent 95% confidence intervals for each cohort. Global multivariable PERMANOVA, adjusted for participant smoking history (pack-years), demonstrated a significant global divergence in microbial community architecture (PERMANOVA *p*=0.004; beta-dispersion *p*=0.004). The concurrent significant beta-dispersion indicates that disease progression is characterised not only by compositional shifts, but also by increased microbial variability (destabilisation) compared to the tightly clustered Healthy baseline. Subsequent stratified post-hoc pairwise PERMANOVA with Benjamini-Hochberg false discovery rate (FDR) correction confirmed significant differences between Healthy and Early-Stage BWs (FDR *p*=0.029), Healthy and Late-Stage BWs (FDR *p*=0.029), and Early-Stage and Late-Stage BWs FDR *p*=0.018). (B) Boxplots displaying the Shannon diversity indices of healthy and lung cancer BWs. Global microbiome stability, assessed using the Kruskal-Wallis omnibus test, identified significant ecological divergence (Shannon *p*=0.014) between cohorts. Subsequent post-hoc pairwise Wilcoxon rank-sum tests identified significantly lower α diversity in Early-Stage BWs compared with Late-Stage BWs. The solid black lines represent the median, with boxes spanning the interquartile range.

Examination of diversity metrics associated with anatomical site (i.e. contralateral vs. TME BWs) using multivariable PERMANOVA revealed significant overarching differences across both the three-group anatomical model (*p*=0.04; **Figure S3A**) and the five-group stage model (*p*=0.03; **Figure 4A**). However, beta-dispersion testing revealed contrasting underlying dynamics: the stage model’s global significance was largely driven by broad ecological destabilisation across the diseased airway (beta dispersion *p*=0.029), whereas the basic anatomical model reflected a genuine compositional shift with equal variance across groups (beta-dispersion *p*=0.743). Despite these overarching structural differences, subsequent stratified post-hoc testing failed to identify distinct microbial community architectures between any specific sites or stages after FDR correction (FDR *p*>0.05). Likewise, Kruskal-Wallis omnibus testing revealed no significant differences in overall α diversity across anatomical sites (*p*=0.70; **Figure S3B**) or disease stages (*p*=0.066; **Figure 4B**). Furthermore, multivariable MaAsLin3 modelling identified zero differentially abundant/prevalent taxa between early-or late-stage disease across both global and anatomical site analyses (FDR *p*>0.1). These results indicate that lung cancer airways exhibit significant dysbiosis, even at early disease stages, and this dysbiosis continues to worsen during the transition to late-stage disease. However, upon anatomical stratification (i.e. contralateral vs. TME), statistical signal is lost, indicating broad impacts across lung cancer airways rather than just at the TME.

**Figure 4.**
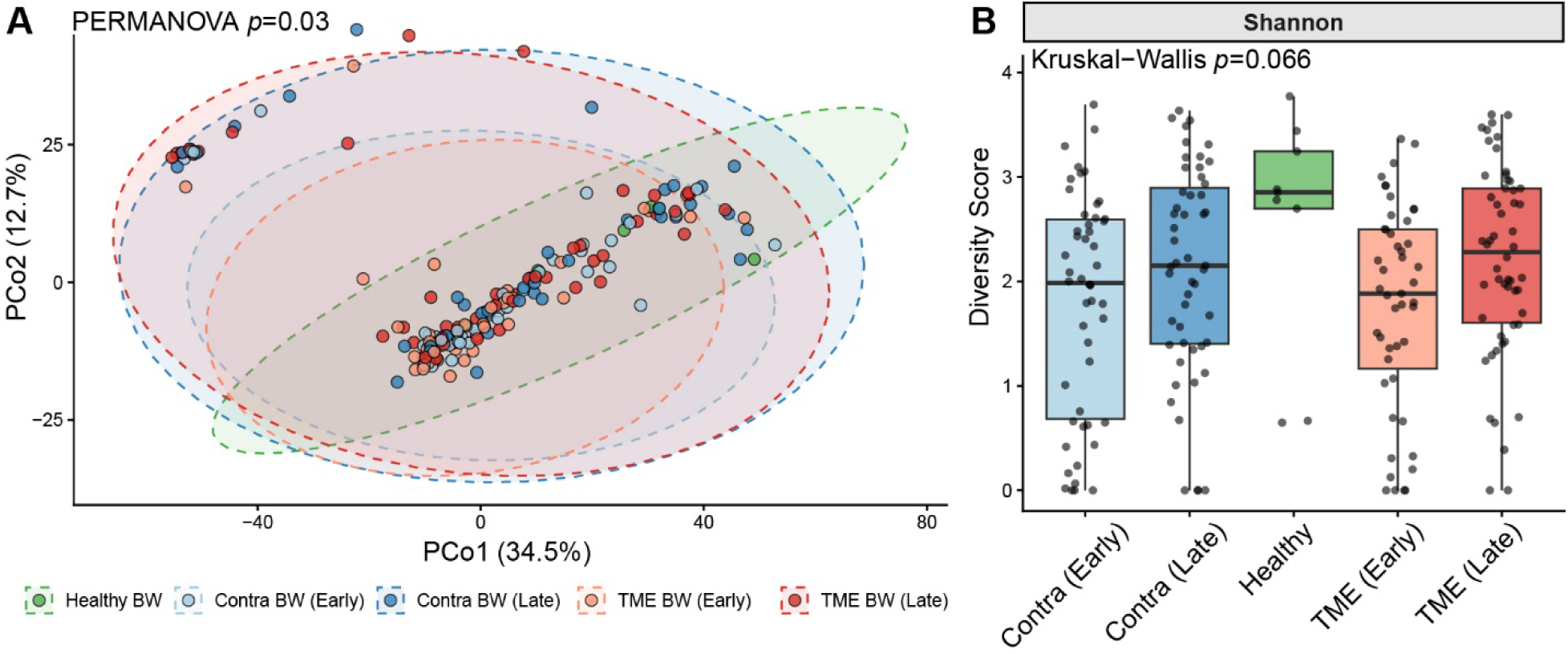
Lung cancer is associated with global structural destabilisation across both local and distant lower airway sites. (A) Principal Coordinate Analysis based on Aitchison distance was used to visualise lung microbiome β diversity in healthy bronchial washes (BWs) and paired contralateral (Contra) and tumour microenvironment (TME) BWs at both early- and late-stage disease. Ellipses represent 95% confidence intervals for each cohort. Global multivariable PERMANOVA, adjusted for participant smoking history (pack-years), demonstrated a significant global divergence in microbial community architecture (PERMANOVA *p*=0.03; beta-dispersion *p*=0.029). The concurrent significant beta-dispersion indicates that this global divergence is characterised not only by compositional shifts, but also by increased microbial variability (destabilisation) across disease sites and stages compared to the tightly clustered Healthy baseline. Consequently, subsequent stratified post-hoc pairwise PERMANOVA with Benjamini-Hochberg false discovery rate (FDR) correction did not identify significant differences between specific sub-cohorts (all FDR *p*>0.05). (B) Boxplots displaying the Shannon diversity indices of healthy vs. paired Contra and TME BWs, stratified by early and late disease stages. Global microbiome stability, assessed using the Kruskal-Wallis omnibus test, identified no significant ecological divergence (Shannon *p*=0.066) between any cohort. The solid black lines represent the median, with boxes spanning the interquartile range.

### Microbiota anatomical gradients from the oral cavity to the lower airways

To evaluate whether the observed lung microbiome signatures were driven by true localised colonisation rather than passive oral microaspiration or bronchoscope contamination, paired multivariable models were used to measure the ecological divergence of lower airway communities (healthy BWs, TME BWs, and contralateral BWs) relative to matched oral washes. Consistent with the strong overall correlation between contralateral and TME BWs (Pearson’s *r* correlation=0.986), our site-specific modelling revealed a shared cancer airway dysbiosis signature that was distinct from healthy controls. Specifically, five core oral commensal taxa (*Streptococcus sanguinis*, *Streptococcus gordonii*, *Streptococcus oralis*, *Veillonella parvula*, and *Actinomyces naeslundii*) were significantly depleted in both TME and contralateral airways relative to the oral cavity and healthy airways, and eight taxa, including *Streptococcus pseudopneumoniae*, *Pseudozyma thailandica*, and *Candidatus* Nanosynbacter sp. HMT−352, were significantly enriched (**Figure 5**).

**Figure 5.**
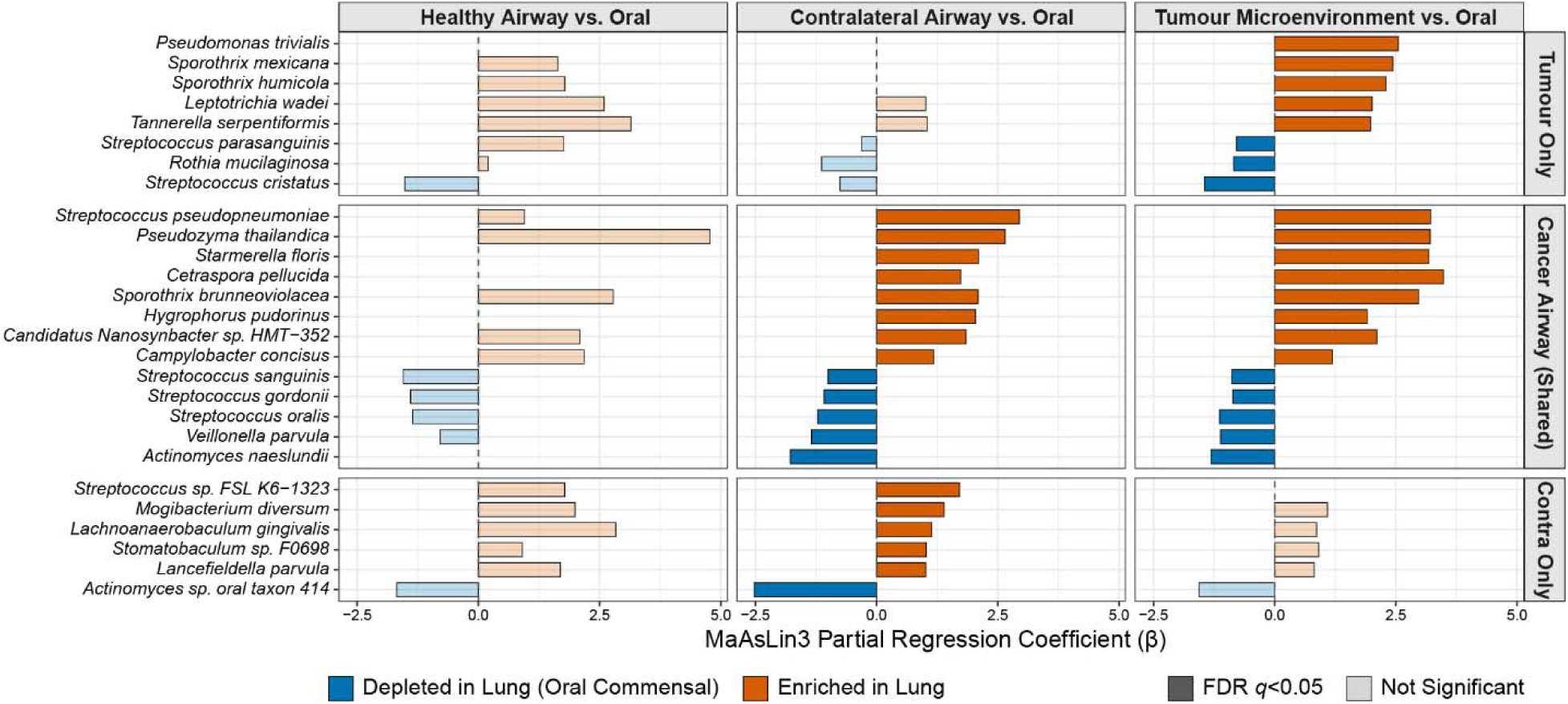
Microbial anatomical gradient between the healthy oral cavity and lower airways in lung cancer and healthy participants. Multivariable mixed-effects modelling (MaAsLin3), controlling for smoking pack-years and biological sex, demonstrated significant enrichment (dark orange) and depletion (dark blue) of microbial taxa within the tumour microenvironment (TME) and the contralateral airway relative to the participant’s matched oral cavity baseline. Both distal lung cancer sites shared strong microbiota commonalities (Pearson’s *r* correlation: 0.986), characterised by the depletion of five core oral commensals (*Streptococcus sanguinis*, *S. gordonii*, *S. oralis*, *Veillonella parvula*, *Actinomyces naeslundii*), and enrichment of eight taxa, including five fungal species. Five taxa were uniquely enriched within the TME, including three non-commensal taxa (*Pseudomonas trivialis* and two *Sporothrix* species), whereas three commensal taxa (*Streptococcus parasanguinis*, *Rothia mucilaginosa*, *Streptococcus cristatus*) were uniquely depleted within the TME. The inclusion of a healthy cohort baseline (left) confirms these ecological shifts are disease-specific phenomena rather than universal pan-airway gradients. Solid bars denote statistical significance (FDR-adjusted *q*<0.05), whereas faded bars represent non-significant fluctuations.

Furthermore, this comparative modelling revealed key microbiota differences unique to specific lung cancer sites. Five microbes were uniquely enriched at the TME, including three non-commensal taxa (*Pseudomonas trivialis* [β coefficient=2.55] and two *Sporothrix* species [β coefficients=2.44 and 2.30], along with two commensal oral taxa (*Leptotrichia wadei* [β coefficient=2.01] and *Tannerella serpentiformis* [β coefficient=1.98]). Three commensal taxa (*S. parasanguinis* [β coefficient=-1.21], *Streptococcus cristatus* [β coefficient=-2.23], and *R. mucilaginosa* [β coefficient= −1.21]) were uniquely depleted at the TME. In contrast, five commensal taxa (*Streptococcus* sp. FSL K6-1323, *M. diversum*, *Lachnoanaerobaculum gingivalis*, *Stomatobaculum* sp. F0698, *Lancefieldella parvula*) were uniquely enriched in contralateral airways, and one (*Actinomyces* sp. oral taxon 414) was uniquely depleted (**Figure 5**). Importantly, none of these anatomical site-specific taxonomic shifts were associated with smoking pack-years or biological sex.

### Lower airway microbial signatures are not associated with lung cancer subtype

To determine whether tumour histological subtypes are associated with specialised lower airway microbial communities, we evaluated LUAD, LUSC, SCLC, and NSCLC-NOS BWs using MaAsLin3 modelling. After controlling for lifetime smoking exposure, we found zero differentially abundant taxa based on subtype (FDR *q*>0.05). Instead, our model confirmed our earlier finding that cumulative smoking exposure exerts significant pressures on the TME microbiome (e.g. *M. diversum* depletion; *q*=0.016). In addition, we found no β (**Figure S4A**; *p*>0.05) or α (**Figure S4B**; *p*>0.05) diversity differences among subtypes. Together, these findings demonstrate that TME microbial composition is shaped primarily by exogenous environmental exposures (such as smoking), along with localised tumour presence, rather than by tumour histology.

### People with LUAD, but not other subtypes, have altered oral microbial architecture (β diversity)

To determine whether tumour histological subtypes are associated with certain oral microbial biomarkers, we compared the oral microbiota of healthy controls and lung cancer participants. Global analysis revealed significant β (**Figure S5A**; PERMANOVA *p*=0.031), but not α (**Figure S5B**), diversity differences across the cohorts. Notably, beta-dispersion testing confirmed that this global divergence was characterised by heterogeneous variance across the cohorts (beta-dispersion *p*=0.046), suggesting that different histological subtypes are associated with varying degrees of ecological destabilisation within the oral microbiome. Post-hoc pairwise PERMANOVA identified that this β diversity variance was driven by the LUAD cohort (FDR *p*=0.020 and FDR *p*=0.050 compared with healthy controls and LUSC participants, respectively), suggesting a histology-specific oral dysbiosis signature. However, using mixed-variable linear models adjusted for smoking pack-years, no individual taxa were robust enough to survive multiple-testing penalties across each lung cancer subtype relative to healthy controls (FDR *q*>0.05), mirroring the lower airway findings (**Figure S4**). Evaluation of the unadjusted data revealed marginal shifts across 22 distinct taxa (raw *p*<0.05), including enrichments in *Schaalia* and *Neisseria* species. Together, these multivariate and univariate results suggest that oral dysbiosis in LUAD is driven by broad, low-amplitude ecological restructuring rather than the dominance of specific univariate biomarkers.

### Early-stage tumour BWs, and late-stage oral washes, exhibit significantly lower cultivability using conventional culture

Comparison of chocolate agar growth across all tested specimens (streak plate score: 0-6) identified significantly lower microbial load in early-stage TME BWs compared with the matched early-stage contralateral BWs (**Figure 6**; *p*<0.001). Furthermore, this early-stage TME suppression trended lower than late-stage TME BWs, late-stage contralateral BWs, and healthy airways (all trending at FDR-adjusted *p*=0.055) pointing toward a critical, transient period of commensal suppression during early tumourigenesis. A trend towards lower cultivability was also observed in late-stage oral washes compared with healthy oral washes (FDR-adjusted *p*=0.055; **Figure 6**).

**Figure 6.**
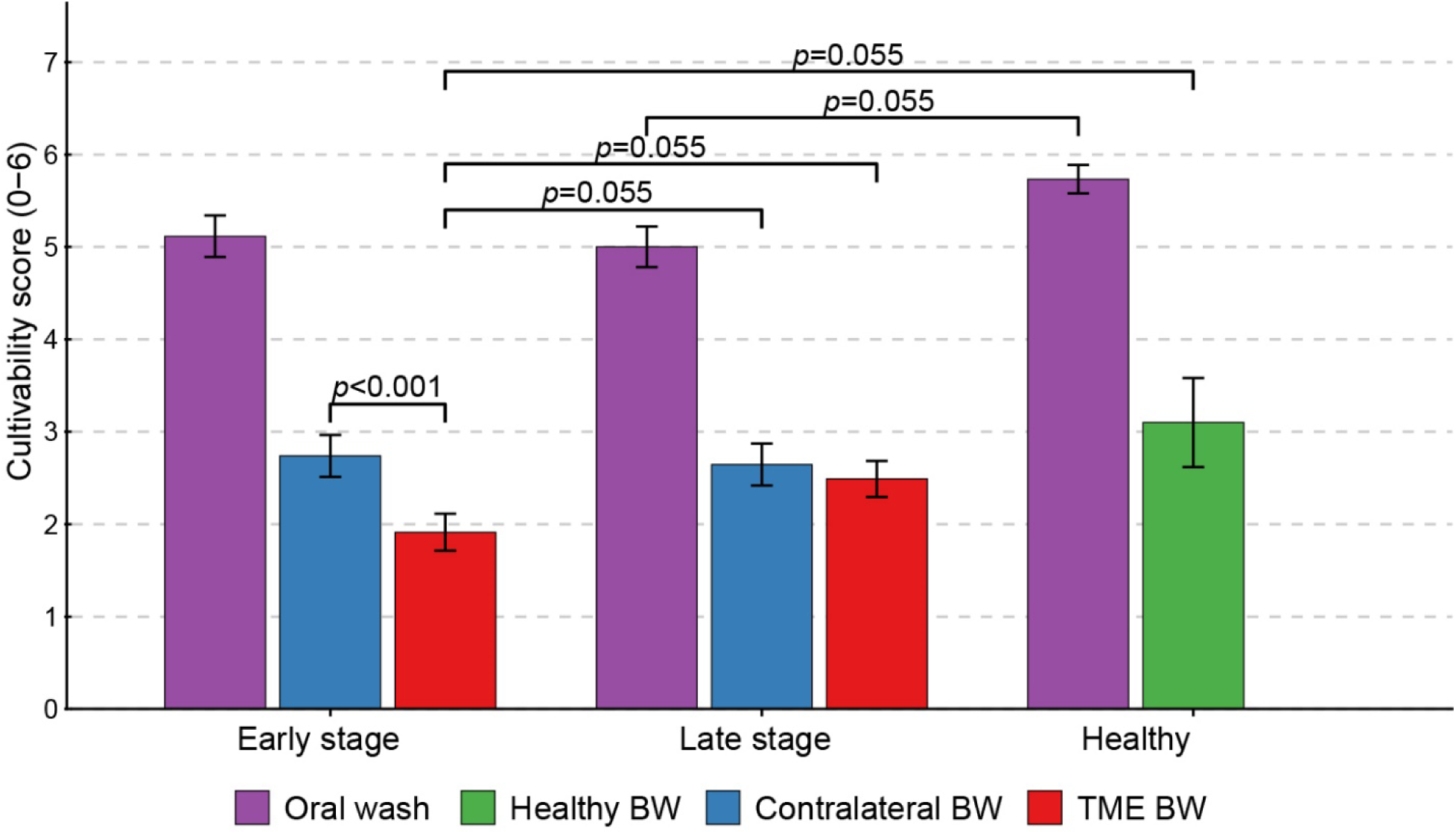
Microbial cultivability of lung and oral washes from people with lung cancer, and healthy controls, using chocolate agar. Semi-quantitative growth scores (0–6) were compared across groups using the Wilcoxon rank-sum test for unpaired comparisons and the Wilcoxon signed-rank test for paired within-cohort comparisons (i.e., matched early-stage tumour microenvironment [TME] vs. early-stage contralateral bronchial washes [BWs]). All *p* values were adjusted for multiple comparisons using the Benjamini-Hochberg false discovery rate correction. Cultivable growth was significantly lower in the early-stage (Stage I-II) TME BWs when compared to matched early-stage contralateral BWs (*p*<0.001). Furthermore, early-stage TME cultivability trended lower when compared to late-stage (Stage III-IV) contralateral BWs (*p*=0.055), late-stage TME BWs (*p*=0.055), and Healthy BWs (*p*=0.055). A similar downward trend was observed in late-stage oral washes compared to Healthy oral washes (*p*=0.055).

Comparison of filamentous fungal culture positivity rates across cohorts identified significantly higher cultivability in contralateral BWs (27/90 [30%] tested specimens) when compared with TME BWs (14/89 [15.7%] tested specimens; *p*<0.01). However, when compared with healthy BWs (2/10 [20%] tested specimens), no significant difference in filamentous fungal culture positivity was seen in either contralateral or TME BWs (both *p*>0.05). Furthermore, no significant differences in filamentous fungus positivity were seen between early-vs. late-stage disease, or when comparing lung cancer subtypes.

### P. trivialis comparative genomics identifies several potentially new species in lung cancer airways

To further investigate *P. trivialis* enrichment in TME BWs (**Figure 5**), we undertook comparative genomic analysis of all *Pseudomonas*-positive metagenomes to better understand the origin of this enriched signal. Due to heavy kitome signal for most pseudomonad hits, including for *P. aeruginosa*, *P. azotoformans*, *P. lactis*, *P. rhodesiae*, and *P. fluorescens*, these taxa were stripped from our metagenome sequencing reads prior to statistical analysis. Just eight pseudomonads survived post-kitome filtering: *Pseudomonas atacamensis*, *Pseudomonas moraviensis*, *Pseudomonas* sp. AK181, *Pseudomonas* sp. HN11, *Pseudomonas* sp. HOU2, *Pseudomonas* sp. J380, *Pseudomonas* sp. MYb193, and *P. trivialis*. Using reference-based read mapping of these raw sequences, phylogenomic reconstruction identified no metagenomic matches to *P. trivialis*; instead, we uncovered a diverse array of pseudomonads, including four specimens (SCHI0241-Contra/-Oral/-TME and SCHI0167-Contra) clustering with *Pseudomonas* sp. AC109, two specimens (SCHI0205-Oral and SCHI0211-Oral) clustering with the *P. carnis* clade, one specimens (SCHI0305-Oral) clustering with *Pseudomonas* sp. FSL L8-0168, and one specimens (SCHI0156-Oral) clustering with the *P. azotoformans* clade. Other specimens (e.g. SCHI0167-TME, SCHI0195-Oral/-Contra/-TME, SCHI0224-Oral) resided on clades with no taxon representation, suggesting that they may be novel *P. fluorescens* group (PFG) species (**Figure S6**).

## Discussion

This study delivers several key advances to the lung cancer microbiome field by combining anatomical site-specific metagenomic profiling, stage-stratified ecological modelling, and conventional culture validation. Our most striking and novel finding is the identification of a distinct functional window in early-stage disease: early-stage TME BWs exhibited significantly reduced microbial cultivability compared to all other airway sites, pointing to a profound, transient suppression of commensal growth during early tumourigenesis. Second, by leveraging a anatomically paired design across early- and late-stage cohorts, we demonstrate that lung cancer induces a progressive, systemic ecological destabilisation across the lower airways. Third, our multivariable models successfully untangle complex environmental confounders, revealing that cumulative smoking drives broad commensal depletion (e.g., *N. subflava*, *G. sanguinis*, *R. mucilaginosa*), and *M. diversum* depletion is uniquely governed by a synergy between heavy smoking and localised tumour presence. Beyond these novel biological insights, our workflow incorporates rigorous negative controls and multimodal culture-independent and culture-dependent validation, providing a robust framework for future biomarker discovery.

It is well established that tobacco smoking alters the lower airway microbiome^37,38^. Given the high tobacco smoking burden in lung cancer cohorts, some prior lung cancer microbiome studies have factored in this covariate by focusing on non-smoker cohorts^14,21,39,40^, or by including smoking status^7,15,18,19,41–44^ or pack-years^11,43^ in statistical models. For example, *Acidovorax* abundance was reportedly increased in the lung tumours of smokers with *TP53*-mutated LUSC^11^, while *Staphylococcus haemolyticus*, *H. influenzae*, *Mycobacterium kansasii*, *S. parasanguinis*, and *Atopobium parvulum* were enriched in smokers with lung cancer^15,18^. Conversely, healthy baseline bacteria such as *Mogibacterium* were reportedly depleted due to tobacco exposure^44^. Furthermore, bacteria within the lung tumours of current smokers are functionally enriched for metabolic pathways capable of degrading cigarette smoke chemicals such as nicotine, toluene, and phenol^20^, suggesting that tobacco exposure actively selects for microbial communities based on their metabolic utility^11,20^.

As our study recruited predominantly current or former smokers (95.3% of the lung cancer cohort, 60% of the healthy cohort; **Table 1**), we controlled for cumulative smoking pack-years using multivariable mixed-effects modelling. This continuous gradient approach revealed that cumulative smoking exposure is an independent ecological driver in lower airways, causing significant, dose-dependent depletion of *N. subflava*, *Gemella sanguinis*, and *R. mucilaginosa* (**Figure 2**), alongside highly localised depletion of *M. diversum* strictly within the TME of heavier smokers (**Figure S1**). The *M. diversum* finding, also reported by others^44^, suggests that this bacterium is uniquely susceptible to the combination of tobacco smoke and localised cancer-driven inflammation. Supporting this hypothesis, our anatomical site-specific modelling revealed that *M. diversum* is uniquely enriched in contralateral airways relative to the oral cavity, but fails to establish similar enrichment within the TME or healthy airways (**Figure 5**). Together, these data suggest that *M. diversum* opportunistically colonises the broader cancer airway but is acutely vulnerable to the toxic microenvironment generated by heavy smoking at the tumour site.

In contrast to commensal depletion, *S. mitis* showed a strong positive correlation with abundance (β=+1.05) in heavier smokers, although significance fell just outside our strict FDR boundary (*q*=0.055). Key oral bacteria, including *S. mitis*, can induce lower airway dysbiosis by upregulating T-helper cell type 17 response^45^, driving increased lower airway inflammation^46^ and fostering a pro-tumourigenic environment^42^. Consistent with this pathology, *S. mitis* enrichment has been reported in lung cancer BALFs compared with benign lung disease^17,18^. Whilst mirroring prior categorical descriptions of *Streptococcus* enrichment and *Neisseria* depletion in smoker airways^37^, our continuous model demonstrates that these shifts are progressive, cumulative events. Taken together, we found that tobacco exposure erodes protective commensal barriers and promotes *S. mitis* proliferation, chronic inflammation, and potential tumourigenesis. This finding raises the intriguing possibility of targeted microbiome restoration following smoking cessation^37^ using *N. subflava*, *G. sanguinis*, and *R. mucilaginosa* to reduce lung cancer risk. Furthermore, the depletion of these specific taxa is noteworthy given that they are core oral commensals; recent evidence similarly highlights the critical role of oral-derived microbiota in shaping the NSCLC immune microenvironment^47^. Taken together, our continuous model demonstrates that tobacco exposure progressively erodes these protective, oral-derived commensal barriers. Crucially, our findings caution against proposing *Streptococcus* as a standalone lung cancer biomarker^42,48,49^, as this signature may simply reflect high cumulative smoking exposure.

α and β diversity are routinely reported in lung cancer microbiome studies; however, different metrics and analysis methods make cross-study consolidation difficult. Nevertheless, global comparisons show a clear trend towards significant β diversity differences between lung cancer and non-malignant lower airway specimens^9,15,17,41,48^. In line with these prior findings, our anatomically paired β diversity analysis revealed that the lung cancer microbiome undergoes profound global species compositional restructuring distinct from healthy controls (PERMANOVA *p*=0.022; **Figure**l**S2A**), which we confirmed to be a true structural shift unconfounded by unequal variance (beta-dispersion *p*=0.678). On a more granular level, we also observed significant community composition divergence between healthy and lung cancer airways when stratified into TME and contralateral BWs (PERMANOVA *p*=0.04; **Figure S3A**). As this anatomical model also exhibited equivalent within-group variance (beta-dispersion *p*=0.743), findings reflect a genuine shift in core community composition rather than transient destabilisation. However, this signal lost significance upon post-hoc pairwise analysis, further supporting the observation that the lower airway undergoes generalised structural microbiome alterations rather than developing localised, site-specific divergence. In contrast, although some studies report increased^7,11,50^ or decreased^15,18,19,43,44^ α diversity in lung cancer, we did not observe global (**Figure S2B**) or anatomically-stratified (**Figure S3B**) α diversity differences in our dataset, in agreement with other studies^9,13,37^. Therefore, when assessing lung cancer airways purely by disease presence or anatomical site, microbiome disruption is characterised by structural compositional shifts (β diversity) rather than changes in community richness or evenness (α diversity).

Whilst global comparisons are informative, lung cancer biomarker discovery requires a thorough understanding of whether microbiome alterations represent a true disease signature versus an association with clinical covariates, such as disease stage. However, just two published studies^18,42^ have compared early- and late-stage lower-airway microbiomes in people with lung cancer. This literature paucity is likely due to the dominance of late-stage lung cancer diagnoses^44,50^, which limits statistical power. Other studies lack tumour staging data^9,16^, prohibiting stage-specific analyses. In the first disease stage-stratified study, Tsay and colleagues^42^ found significant β, but not α, diversity differences in their NSCLC cohort. Interestingly, they noted an increased abundance of oral taxa, including *Veillonella, Streptococcus, Prevotella, and Rothia*, in latel71stage NSCLC. They also found that lower airway microbiota in late-stage participants was compositionally more similar to the buccal microbiota than in early-stage participants, suggesting that late-stage disease may be associated with a greater influx or retention of oral microbes, potentially facilitated by a progressive breakdown in localised mucosal immunity. In the second study^18^, a comparison of preinvasive LUAD *in situ*, minimally invasive LUAD, and invasive LUAD cohorts found no significant α or β differences, although the authors identified specific individual taxa with altered abundance in more invasive disease.

Like Tsay and colleagues, we identified significant global β diversity differences across healthy, early-stage, and late-stage cohorts (PERMANOVA *p*=0.004; **Figure 3A**). Upon cohort stratification, we confirmed that early- and late-stage BWs maintained significant β diversity differences (Healthy vs. Early-Stage BWs: FDR *p*=0.029; Healthy vs. Late-Stage BWs: FDR *p*=0.029; Early-Stage vs. Late-Stage BWs: FDR *p*=0.018). Interestingly, these pairwise microbial composition differences lost significance when stratified by lung anatomical site and stage (**Figure 4**) or just anatomical site (**Figure S3**). This widespread loss of localised significance, paired with significant overarching beta-dispersion (*p*=0.029), suggests that lung cancer induces generalised ecological destabilisation throughout the entire lower airway, rather than generating a localised tumour microbiome signature.

However, unlike Tsay *et al.*, we found that disease stage significantly impacts α diversity. Omnibus testing revealed significant overarching α diversity differences across staged cohorts, driven specifically by much lower α diversity in early-stage BWs compared with late-stage BWs (FDR *p*=0.012; **Figure 3B**). Also dissimilar to the prior studies, we failed to identify any significantly enriched or depleted taxa strictly associated with disease stage using our MaAsLin3 model, even with a more relaxed FDR cut-off of 0.1 and the removal of biological sex as a covariate. Collectively, our results strongly indicate that lung cancer drives substantial site-specific restructuring and ecological destabilisation (β diversity) of the lower airway microbiome compared with healthy airways, a disruption that begins early and persists throughout disease progression. Conversely, α diversity alterations are strictly tied to disease progression, reflecting a transient collapse in community diversity during early tumourigenesis that recovers by late-stage disease.

Controlling for the oral microbiome and healthy baselines, analysis of TME and contralateral BWs revealed significant site-specific microbial differences. The TME was uniquely enriched in *Sporothrix mexicana*, *Sporothrix humicola*, *L. wadei*, *T. serpentiformis*, and a PFG signature assigned to *P. trivialis*. In contrast, the contralateral airway was uniquely enriched in *Streptococcus* sp. FSL K6-1323, *M. diversum*, *L. gingivalis*, *Stomatobaculum* sp. F0698, and *L. parvula* (**Figure 5**). Furthermore, we identified eight significantly enriched taxa (*S. pseudopneumoniae*, *P. thailandica*, *Starmerella floris*, *Cetraspora pellucida*, *Sporothrix brunneoviolacea*, *Hygrophorus pudorinus*, *C.* Nansynbacter sp. HMT-352, *Campylobacter concisus*), and five significantly depleted taxa (*S. sanguinis*, *S. gordonii*, *S. oralis*, *V. parvula*, *A. naeslundii*), across lung cancer airways when compared with healthy controls (**Figure 5**). *T. serpentiformis* and *L. wadei* are periodontitis-associated anaerobes that we postulate may thrive in the low-oxygen, nutrient-rich necrotic TME, which mimics their native gum disease environment, whereas PFG species and *Sporothrix* spp. are ubiquitous plant-, soil-, or wood-associated microbes that are constantly inhaled, potentially via aerosolised cigarette tobacco. Although tumour-associated architectural distortion, airway obstruction, and cilia paralysis may enable these transient environmental microbes to accumulate and fester within the localised TME, we acknowledge the shortcomings of relative abundance profiling. Given our culture data demonstrated significantly reduced absolute microbial load within the early-stage TME (**Figure 6**), the apparent enrichment of these ubiquitous environmental taxa may potentially reflect kitome inflation in low-biomass specimens.

Conversely, we observed significant *S. parasanguinis*, *R. mucilaginosa*, and *S. cristatus* depletion uniquely at the TME, and significant *Actinomyces* sp. oral taxon 414 depletion uniquely at the contralateral site (**Figure 5**). Although previous studies have reported enrichment of other pathogenic or opportunistic taxa within the TME, such as *Veillonella, Megasphaera, Bradyrhizobium japonicum, Capnocytophaga, Sediminibacterium, Gemella, Blautia*, and *Oscillispira*^7,15,16^, these differences likely reflect broader geographical and environmental influences on lung microbial ecology. Nonetheless, a consistent pattern is evident across studies: that is, that lung cancer drives the depletion of protective commensals alongside the selective enrichment of opportunistically pathogenic taxa. Collectively, our findings consolidate prior work showing that the TME fosters a distinct microbial niche that is independent of oral cavity topology and the contralateral lung.

Culture-independent sequencing approaches cannot readily detect absolute microbial load differences due to their inherent compositional data format. As these technologies measure relative proportions rather than absolute numbers, a massive physical bloom of one taxon mathematically forces the apparent decline of others, masking true biomass fluctuations. Integrating conventional culture into culture-independent investigations therefore provides valuable orthogonal validation by quantifying cultivable microbial load across specimen types. Furthermore, conventional culture distinguishes organisms that are metabolically active and capable of shaping the TME from those merely detectable as remnant DNA. Despite these advantages, just one prior study has integrated culture-dependent and -independent techniques. In their 2023 study, Sun and colleagues undertook culturomics alongside 16S rRNA metataxonomics to compare and confirm lung and oral microbiota^51^. They isolated 20 bacteria from BALF specimens from at least half of their lung cancer cohort, noting that *Streptococcus* and *Veillonella* were highly dominant, *Prevotella* and *Veillonella* decreased from the oral cavity to the lung, *Pseudomonas* increased, and *G. sanguinis* and *Streptococcus intermedius* were isolated exclusively from NSCLC. Culturing viable microbes directly from lung cancer specimens is critically important for understanding how specific organisms and strains actively contribute to local inflammation, modulate tumourl71associated immune responses, or influence tumour proliferation pathways.

Although our study did not undertake culturomics, our semi-quantitative agar scoring approach crucially identified significantly lower cultivable microbial growth in the early-stage TME compared with all other BW types (**Figure 6**). This striking finding points to a critical period in early lung tumourigenesis, during which the TME transiently, yet profoundly, suppresses commensal microbial growth. This localised suppression may reflect a hyperactive mucosal immune response: during early tumour development, the TME is characterised by an influx of macrophages and neutrophils, and the production of reactive oxygen species, thus creating a hostile zone for commensal microbes^52^. Another possibility is the targeted secretion of antimicrobial peptides by the developing tumour, such as human β-defensins (e.g. hBD-1)^53^. Our observation offers unique opportunities for early-stage biomarker discovery and the development of new therapeutic strategies aimed at restoring a more favourable TME microbiome profile at a potentially critical juncture in lung cancer pathogenesis, and deserves further exploration.

Current literature is split on whether different histological subtypes are associated with microbiome alterations, although most studies have reported no significant α or β diversity differences^9–11,17,39,42,48,54–56^. Similarly, we observed no significant α or β diversity differences, or differentially abundant taxa, when comparing LUAD, LUSC, SCLC, and NSCLC-NOS (**Figure S4**). Although most prior lung cancer microbiome studies have not undertaken subtypel71specific comparisons, our findings diverge from those of Greathouse and colleagues, who reported *Acidovorax, Klebsiella, Rhodoferax,* and *Anaerococcus* enrichment in LUSC relative to LUAD tissue^11^; however, a key distinction is that Greathouse and colleagues analysed tumour tissue. Nevertheless, these contrasting observations underscore the need for future studies that directly interrogate tumourl71specific microbial signatures across histological and molecular subtypes, particularly in the context of increased genomic subclassification and precisionl71based oncology^57^. Our findings suggest that the extratumoural lung microbiome is not shaped by tumour histology; however, larger sample sizes, particularly for SCLC and rarer histological subtypes, would be needed for definitive confirmation. Furthermore, unlike studies examining intratumoural tissue specimens, our analysis focused on extratumoural BWs, which may capture a broader airway community less acutely bound to local histological niches. Although not explored in this study, functional profiling of lung cancer subtypes using host-microbe epigenomics, proteomics, or metabolomics may uncover biomarkers that are strongly linked to subtype.

Oral microbial signatures have been proposed as an appealing, nonl71invasive biospecimen for lung cancer biomarker development, with several studies reporting taxonomic and oral microbiome compositional differences in lung cancer cohorts^39,48,49,58,59^. In our cohort, global comparisons revealed significant overarching structural divergence in the oral microbiome across lung cancer subtypes (PERMANOVA *p*=0.031; **Figure S5A**), accompanied by significant ecological destabilisation (beta-dispersion *p*=0.046), indicating that lung cancer presence disrupts the oral microbiome compared with the healthy baseline. Upon pairwise testing, it became clear that this global divergence was primarily driven by the LUAD cohort, as we observed significant β diversity differences between LUAD and healthy oral microbiota (FDR *p*=0.020), with a borderline difference between LUAD and LUSC (FDR *p*=0.050). Despite this significant β diversity shift, we found no strong evidence that tumour-associated microbial patterns originate from oral cavity dysbiosis, and, reflective of the broad ecological destabilisation seen in LUAD, no taxa were consistently enriched or depleted in any subtype. Instead, our findings support the hypothesis that distinct lung cancer-associated microbial signatures arise from local ecological selection pressures within the lower airways, aligning with prior work demonstrating clear divergence between TME and oral microbiota^48,59^. Taken together, our results indicate that further investigation of oral β diversity disturbances in LUAD cohorts is warranted; however, they temper enthusiasm for the use of oral sampling as a non-invasive lung cancer biomarker for other histological subtypes.

Our finding of unique *P. trivialis* enrichment in TME BWs (**Figure 5**) prompted us to further investigate this fluorescent, grass-dwelling bacterium^60^ in our metagenomic reads using comparative genomics. Despite this significant signal, none of our metagenomes possessed *P. trivialis* according to phylogenomic reconstruction; instead, a wide array of PFG taxa, including potentially novel species, were identified (**Figure S6**). Whilst an incorrect taxonomic assignment by Kraken2, our *P. trivialis* finding nevertheless suggests that PFG bacteria may play an outsized role in the TME microbiome. PFG species are rarely associated with human infections, typically affecting only immunocompromised people, but are common in soil and water, and can colonise human airways, skin, and the gastrointestinal tract. Given that cancer patients receiving treatment frequently become immunocompromised, PFG infection may represent an unrecognised risk for this cohort. As many PFG species likely cannot grow on MacConkey agar, we overlooked these taxa in our conventional culture-based assessments. Future work will involve isolation and whole-genome sequencing of these organisms from lung cancer specimens, which in turn, will facilitate improvements in Kraken2 and other taxonomy-based bioinformatic tools.

Strengths of our study include its prospective design and our near-universal collection of paired tumour and contralateral BWs, alongside matched oral washes (*n*=92; 86.8% of participants), which permitted precise mixed-effects modelling that inherently controlled for intra-patient variability and topographical contamination. However, we acknowledge several limitations. Our lung cancer cohort possessed a substantially higher mean smoking history (31.8±16.5 pack-years) compared with healthy controls (3.3±2.3 pack-years), reflecting the inherent clinical difficulty in recruiting heavy-smoking healthy volunteers without underlying respiratory disease. Although statistically adjusted, fully decoupling the smoking and lung cancer microbiomes requires longitudinally tracking heavy smokers who do not develop lung cancer. Reliance on short-read sequencing limited taxonomic resolution, and bioinformatic decontamination was required to remove microbial DNA introduced by reagent and kit components, which potentially included real signal in our BWs. Future low-biomass studies should prioritise certified DNA-free reagents to optimise signal-to-noise ratios and should integrate long-read sequencing to enhance species-level assignments. Sampling was restricted to the extratumoural space; incorporating matched endobronchial/transbronchial biopsies or surgical resections is needed to confirm whether these signatures persist within the intratumoural niche. We did not undertake selective culture and whole-genome sequencing of key taxa (e.g. PFG species, *N. subflava*, *M. diversum*, *P. thailandica*, *Sporothrix* spp.) to confirm their presence in our clinical specimens. The absence of matched blood and stool specimens precluded the evaluation of less invasive circulatory or faecal biomarkers. The addition of RNA-seq^11,14,21,41–43,61^, host-depleted metatranscriptomics, methylomics, proteomics, and metabolomics^17,62^ would have enabled the characterisation of microbial-host transcriptional activity, epigenomic fingerprints, and the detection of RNA viruses and metabolic by-products. As these multi-omics technologies become more accessible, their integration will be essential to unravel the complex microbe-microbe and microbe-host oncogenic pathways underpinning lung cancer development and progression.

## Conclusions and future perspectives

By combining high-resolution shotgun metagenomics, stage-stratified anatomical site-specific modelling, and orthogonal culture-based validation, this study provides a comprehensive characterisation of the extratumoural lung cancer microbiome. Our findings demonstrate that lung cancer induces a systemic, progressive ecological destabilisation of the lower airways that begins early in disease pathogenesis, marked by a striking, transient suppression of commensal cultivability within the early-stage TME. Furthermore, by rigorously controlling for cumulative tobacco smoking, we show that lower airway dysbiosis is driven by a complex interplay of environmental exposures and localised tumour pressures, independent of histological subtype. These insights advance our mechanistic understanding of lung cancer-associated microbial ecology and underscore the potential of integrating microbial viability metrics into future diagnostic and therapeutic frameworks. Future investigations should expand upon these findings by incorporating longitudinal sampling and diverse global cohorts (younger adults, older women, el71cigarette users, nonl71smokers, individuals undergoing immunotherapy, radiotherapy, chemotherapy, or surgical resection, and populations from underl71studied regions, especially Africa, the Middle East, and Southern and Southeastern Asia) to capture evolving lung cancer demographics and refine diseasel71specific microbial biomarkers. Investigation of other host-microbe biomarkers, such as methylation signatures, may also unveil new pathogenesis pathways.

## Data Availability

The human- and kitome-depleted metagenomic reads have been deposited under BioProject PRJNA1539961.

## Acknowledgements

We wish to thank Barry Pritchard, Ben Clarke, and Claire Winterbourne (Sunshine Coast Health Institute) for laboratory support, the Queensland Cyber Infrastructure Foundation (University of Queensland) for high-performance computing support, and the Centre for Bioinnovation (Paige Burton, Abigail Elizur) for project support. We acknowledge the use of AI (Google Gemini 3 Series) for manuscript refinement.

