## Supplemental Methods for "The LAMp-LC Study: Unveiling the lung microbiome in Australians with early- and late-stage lung cancer using shotgun metagenomics"

**Tumour staging.** Lung tumours were staged using the Tumour, Node, and Metastasis (TNM) system (9th edition)^1^. Tumour typing and subtyping were undertaken as part of routine Pathology Queensland cytology and histopathology testing. Information regarding final confirmed lung cancer diagnosis, subtype (LUAD, LUSC, NSCLC-NOS, SCLC, and carcinoid) and stage (early stage [I-II] vs late stage [III-IV]) was collated (**Table S1**). Routine Pathology Queensland lung cancer genomic alteration screening was undertaken on respective tumour tissue biopsies to test for *EGFR*, *KRAS*, and *BRAF* mutations, and *ALK*, *ROS1* rearrangements, along with immunohistochemistry for *PD-L1* expression (low; 1-49%; high, ≥50%)^2,3^. At the time of recruitment, *TP53*, *MET*, *RET*, *HER2*, and *PIK3CA* mutations were not routinely examined, as per Pathology Queensland workflows^2^. For some specimens, genomic testing was unable to be undertaken due to insufficient DNA yield. Due to ethical constraints restricting analysis of the human component of our shotgun metagenomes, we were unable to derive tumour genotypes from these data.

**Host DNA depletion and DNA extraction.** Approximately ~500µL specimen was depleted of human DNA using the “Benzonase 1” protocol^4^, followed by resuspension into 200μL lysis buffer (20mM Tris-HCl pH 8.0, 2mM sodium EDTA, and 1.2% Triton X-100 [Sigma-Aldrich, Castle Hill, NSW, Australia]). For 70 specimens, cell lysis was undertaken using an in-house metapolyzyme concoction of lysozyme, chitinase, achromopeptidase, mutanolysin, lysostaphin, and zymolyase, as described elsewhere^5^. For the remaining 250 specimens, an enzyme-free cell lysis protocol was undertaken using ~100μL of 0.1 and 0.5 mm zirconia beads (Daintree Scientific, St Helens, TAS, Australia) and bead beating at 2 x 7500 rpm for 60 sec (with a 30 sec pause in between) in DNase-free 2mL O-ring tubes (SSIBio, Lod, CA, USA) using a Precellys Evolution Touch Homogeniser (Bertin Technologies, Montigny-le-Bretonneux, France). DNA from both enzymatic and bead beating cell lysis protocols was purified using the ‘Gram-positive Bacteria’ protocol of the DNeasy Blood and Tissue kit (QIAGEN, Clayton, VIC, Australia); for the bead beating protocol, purification commenced at the Buffer AL and Proteinase K addition step. Extracted DNA was quantified using a Qubit 4 fluorimeter (Thermo Fisher Scientific, Scoresby, VIC, Australia). Reagent-only controls were included in each metagenomic sequencing batch (**Table S3**) and used to subtract background microbial DNA contamination in extraction and sequencing reagents. These controls were handled in an identical manner to specimens, except for no specimen addition.

**Taxonomic assignment of metagenomic reads.** Following contaminant depletion, we assigned microbial taxonomies against the pre-built Kraken 2 Standard v2/26/2026 (Refseq archaea, bacteria, viral, plasmid, human, UniVec_Core) database. For fungal taxonomy, a custom Fungal Kraken 2 database was constructed using 5,012 NCBI GenBank non-redundant fungal genomes (downloaded 21Apr26), excluding ‘Atypical genomes’ and ‘Metagenome-assembled genomes’. No-template controls (NTCs) sequenced in each run were used to strip out kitome signal (based on Kraken 2 taxonomic assignments) using seqtk, with 5% of the NTC signal for each microbe removed from each specimen within the same sequencing run. This 5% NTC filter was chosen as we found that a higher percentage removed real microbial signal, as determined by qPCR/whole-genome sequencing confirmed culture positivity. Kraken 2 outputs from Standard and Fungal databases were merged prior to statistical analysis. Due to difficulties with creating a Bracken database for our custom Fungal database, we did not further pursue this tool.

As experienced by others^6^, we chose not to include R decontam package filtering, as we found it inadvertently removed a large fraction of biological important signal. Instead, we undertook kitome decontamination using one or more of the following parameters for each taxon: minimum 5000 reads for in at least one clinical specimen (excluding Epstein-Barr virus), ratio of >3 for specimen maximum:NTC maximum, >10000 reads difference between specimen sum and NTC sum, ratio >1 for specimen:NTC 95% median, taxon prevalence >3 in the clinical specimens, taxon prevalence <17 in the 33 NTCs, and ratio >0.2 for specimen:NTC taxon prevalence. Known enzyme constituents (*Streptomyces* [achromopeptidase, mutanolysin], *Arthrobacter* [zymolyase], *Lysobacter* [chitinase]), along with a probable Milli-Q water contaminant, *Cellulosimicrobium*, were bioinformatically removed. We subsequently removed ‘*Mycobacterium canettii*’ and ‘*Coniochaeta pulveracea*’ from our raw count data matrix due to near-perfect read alignment to known kitome contaminants (in the case of *M. canetti*) and very high prevalence across most metagenomes (in the case of *C. pulveracea*), including metagenomes with otherwise no detectable microbiome, suggesting potential non-depleted human read misassignment.

**Available metagenomes.** Eleven specimens (five tumour microenvironment [TME] BWs, five contralateral BWs, one oral wash) failed to yield any detectable DNA and were excluded from metagenomic sequencing. A further 13 specimens yielded no detectable microbiome after metagenomic sequencing and data filtering (**Table S1**). In total, 279 metagenomes from the lung cancer cohort (93, 95, and 91 contralateral, TME, and oral washes, respectively), and 24 metagenomes from the healthy cohort (nine BWs and 15 oral washes), had detectable microbiomes. Paired BWs were available from 88 lung cancer participants, 75 with a matched oral wash.

**Conventional culture and qPCR testing.** As our laboratory has an interest in *Pseudomonas aeruginosa*, *Achromobacter* spp., and *Stenotrophomonas* *maltophilia*, we undertook selective culture and subsequent qPCR testing to confirm the identity of these organisms in lung cancer specimens, as described previously^7-9^. We also glycerol-stocked all non-lactose-fermenting culture growth, and filamentous fungal growth, for future whole-genome sequencing.

***Pseudomonas* speciation*.*** To speciate the Kraken 2-assigned ‘*Pseudomonas trivialis*’ metagenomic reads, we undertook comparative genomic analysis of all ‘*Pseudomonas trivialis*’, ‘*Pseudomonas* sp. AK181’, ‘*Pseudomonas atacamensis*’, ‘*Pseudomonas* sp. MYb193’, ‘*Pseudomonas* sp. HOU2’, ‘*Pseudomonas* sp. HN11’, ‘*Pseudomonas moraviensis*’, and ‘*Pseudomonas* sp. J380’ positive metagenomes using SPANDx v4.3.1^10^. *Pseudomonas* species assemblies *P. aeruginosa* PAO1 (GCF_000006765.1), *Pseudomonas atacamensis* SM1 (GCF_017167965.1), *P. atagonensis* PS14 (RefSeq: GCF_011369485.1), *P. azotoformans* LMG 21611 (GCF_900103345.1), *P. baetica* LMG 25716 (GCF_002813455.1), *P. brenneri* DSM 15294 (GCF_007858285.1), *P. canadensis* GZH03 (GCF_030980095.1), *P. carnis* 20TX0167 (GCF_024722005.1), *P. congelans* DSM 14939 (GCF_900103225.1), *P. crudilactis* UCMA 17988 (GCF_013433315.1), *P. cyclaminis* MAFF 301449 (GCF_015163715.1), *P. fjordensis* 1331593 (GCF_976987115.1), *P. fluorescens* ATCC 13525 (GCF_900215245.1), *P. gorinensis* NT2 (GCF_042015035.1), *P. granadensis* LMG 27940 (GCF_900105485.1), *P. lactis* DSM 29167 (GCF_001439845.1), *P. libanensis* DSM 17149 (GCF_001439685.1), *P. mandelii* LMG 21607 (GCF_900106065.1), *P. mediterranea* DSM 16733 (GCF_900106005.1), *P. moraviensis* B21-039 (GCF_024748715.1), *P. moraviensis* subsp. stanleyae 71 (GCF_036283065.1), *P. mucidolens* NCTC 8068 (GCF_900475945.1), *P. ogarae* SWRI108 (GCF_014268695.2), *P. neuropathica* P155 (GCF_015461835.1), *P. paralactis* DSM 29164 (GCF_001439735.1), *P. pharyngis* BML-PP036 (GCF_021602345.1), *P. rhodesiae* NL2019 (GCF_013285305.1), *P. salmasensis* SWRI126 (GCF_014268375.2), *Pseudomonas* sp. AC109 (GCF_979257885.1), *Pseudomonas* sp. AK181 (GCF_051122975.1), *Pseudomonas* sp. FJ2-5-13 (GCF_029201605.1), *Pseudomonas* sp. FP215 (GCF_030687795.1), *Pseudomonas* sp. FSL L8-0168 (GCF_037976155.1), *Pseudomonas* sp. GXZC (GCF_026967615.1), *Pseudomonas* sp. HN8-3 (GCF_020790195.1), *Pseudomonas* sp. J380 (GCF_009827115.1), *Pseudomonas* sp. MYb193 (GCF_002966775.1), *Pseudomonas* sp. OST1919 (GCF_015277575.1), *P. sputi* BML-PP014 (GCF_021603585.1), *P. synxantha* NCTC 10696 (GCF_901482615.1), *P. tolaasii* NCPPB 2192 (GCF_002813445.1), and *P. trivialis* W04_8 (GCF_036864995.1) were included for comparison using the ‘--assemblies’ flag. *P. lactis* DSM 29167 was used as the alignment reference genome.

**Alpha (α) and beta (β) diversity analyses*.*** Microbial community diversity was evaluated at the species level using the vegan (v2.7-3) package in R^11^*.* Within-sample ecological diversity (α diversity) was quantified using the Shannon diversity index^12^. To accommodate the specific clinical sampling design of this study, non-parametric statistical tests were employed as follows: (1) α diversity differences between paired contralateral and tumour BWs within the same participant were evaluated using the Wilcoxon signed-rank test^13^; (2) α diversity differences between two independent clinical cohorts (e.g., healthy controls vs. all lung cancer participants) were evaluated using the two-sample Wilcoxon rank-sum test^13^; and (3) variance across three or more independent groups (e.g. anatomic site, disease stage) was assessed using the Kruskal-Wallis omnibus tests^14^. Significant omnibus results were followed by pairwise Wilcoxon testing with Benjamini-Hochberg false discovery rate (FDR) correction^15^.

Between-sample microbial composition (β diversity) was evaluated using Aitchison distance, which applies Euclidean distance to centred log-ratio (CLR) transformed count data (using a pseudocount of 1 to adjust for zero values) to handle the compositional nature of microbiome datasets. Structural similarities and community clustering were visualised using Principal Coordinates Analysis (PCoA). Statistical differences in global community composition were assessed using multivariable Permutational Multivariate Analysis of Variance (PERMANOVA)^16^ via the adonis2 function^17^ with 999 permutations. Consistent with the α diversity framework, PERMANOVA designs were tailored to the specific comparative cohorts. For all paired spatial analyses evaluating within-participant divergence, the PERMANOVA was explicitly stratified by participant ID to control for inter-participant baseline microbiome variation. Where appropriate, global multivariable PERMANOVA was followed by stratified post-hoc pairwise PERMANOVA, with resulting *p*-values subjected to Benjamini-Hochberg FDR correction.

**MaAsLin3 analyses*.*** To identify microbial taxa significantly associated with clinical disease state, spatial sampling site, and temporal progression whilst controlling for confounding clinical covariates, multivariable association modelling was performed using MaAsLin3^18^. MaAsLin3 uses a two-part ‘hurdle’ framework to evaluate zero-inflated microbiome data. This approach independently models two distinct ecological metrics for each taxon: prevalence (the probability of presence/absence evaluated via logistic regression) and abundance (evaluated via linear regression of the non-zero counts). To reduce the FDR burden from multiple testing, the input community matrix was filtered to retain only core taxa present at a minimum relative abundance of 0.01% in at least 20% of all specimens. Raw count data subsequently underwent Total Sum Scaling normalisation followed by natural logarithm transformation prior to linear modelling. The ‘small_random_effects=TRUE’ parameter was included to account for small group sizes, such as in the paired models tracking participant IDs.

To accurately capture the diverse ecological dynamics of the cohort, the dataset was evaluated using three distinct modelling strategies, most of which included lifetime smoking exposure (pack-years) as a fixed-effect covariate to isolate disease-specific microbial shifts from environmental tobacco signatures. Smoking exposure was excluded in our stratified early- and late-stage spatial divergence models to preserve degrees of freedom, but was retained in all temporal progression assessments. (1) Global Disease Assessment: A standard multivariable linear model was use to compare healthy controls (baseline reference) against the global lung cancer cohort to identify systemic taxonomic shift; (2) Spatial Divergence (Mixed-Effects): To evaluate localised spatial divergence within the lung cancer cohort, paired mixed-effects models were employed to compare tumour BWs against matched contralateral BWs. In these models, the sampling site was designated as a fixed effect, whilst participant ID was included as a random effect to mathematically control for participant-specific baseline microbiome variation. Spatial divergence was evaluated independently within both early-stage and late-stage cohorts; (3) Temporal Progression and Histology (Stratified Analysis): Additional independent multivariable models were used to evaluate microbial differences associated with disease progression (early vs. late stage) and histological tumour subtypes (e.g., LUAD vs. LUSC).

Demographic covariates such as age and biological sex were initially evaluated as potential confounders across all analyses. However, to prevent model overfitting and to preserve statistical degrees of freedom (power) within the smaller, stratified subsets (e.g., early vs. late cohorts), non-significant demographic covariates were excluded from the final multivariable models. Specifically, age was excluded from all final models due to a lack of significant confounding, whilst sex was retained in the global and contralateral assessments but excluded from highly stratified subsets (e.g., early vs. late cohorts) to preserve statistical power. Statistical significance for all MaAsLin3 models was evaluated using individual *p*-values, which were subsequently adjusted for multiple hypothesis testing using the Benjamini-Hochberg FDR procedure^15^. The resulting FDR-adjusted *p*-values (reported as *q*-values) denote the significance of differential abundance, with a stringent threshold of *q*<0.05 used to define statistically significant taxonomic shifts.

**Culture-based statistical analyses**. Comparison of cultivable chocolate agar growth across different cohorts and disease parameters was conducted using Wilcoxon signed-rank testing for paired intra-participant data, and Wilcoxon rank-sum testing for unpaired, independent samples^13^. Ordinal logistic regression using a Generalised Estimating Equations^19^ model was also used to assess disease state and specimen type clustering and interactions. Kruskal-Wallis omnibus testing was used to examine cultivability differences across lung cancer subtypes^14^. For fungal assessments, filamentous fungal growth was recorded as positive if at least one distinguishable colony was observed after 5-8 days of incubation, irrespective of the specific agar medium. Due to the internally controlled nature of our sampling, McNemar’s test^20^ was used to compare categorical (‘yes/no’) filamentous fungal growth between paired tumour and contralateral washes. Fisher’s exact two-tailed testing^21^ was used for all other comparisons.
