## Supplementary Data for "The LAMp-LC Study: Unveiling the lung microbiome in Australians with early- and late-stage lung cancer using shotgun metagenomics"

**Table S1. Lung cancer (LC) participants, disease stage, collected specimens, and metagenome-sequenced specimens.**

| **Participant** | **Tumour TNM Stage** | **Tumour subtype** | **Smoking Pack-Years** | **Age Range** | **Sex** | **Disease Stage (I-IV)** | **Stage Category** | **Oral Wash** | **BW –Tumour** | **BW – Contra** | **SRA IDs** |
| --- | --- | --- | --- | --- | --- | --- | --- | --- | --- | --- | --- |
| SCHI0034 | T2aN0M0 | LUAD | 30 | 86-90 | M | IB | Early | N | Y | Y | SRR41050908, SRR41050801 |
| SCHI0036 | T1bN2M1c | SCLC | 40 | 61-65 | F | IVB | Late | N | Y | Y | SRR41050790, SRR41050899 |
| SCHI0043 | T4N0M0 | LUAD | 20 | 66-70 | M | IIIA | Late | N | Y | Y | SRR41050888, SRR41050709 |
| SCHI0044 | T1cN2M0 | SCLC | 40 | 66-70 | F | IIIA | Late | N | Y | Y | SRR41050698, SRR41050990 |
| SCHI0049 | T3N0M0 | LUAD | 50 | 81-85 | M | IIA | Early | N | Y | Y | SRR41050979, SRR41050785 |
| SCHI0055 | T4N2M1c | SCLC | 70 | 76-80 | M | IVB | Late | N | Y | Y | SRR41050773, SRR41050882 |
| SCHI0063 | T1bN0M0 | LUAD | 20 | 81-85 | F | IA2 | Early | N | Y | Y | SRR41050871, SRR41050860 |
| SCHI0075 | T1bN0M0 | LUAD | 20 | 76-80 | F | IA2 | Early | N | Y | Y | SRR41050960, SRR41050949 |
| SCHI0079 | T1aN0M0 | LUSC | 30 | 66-70 | F | IAI | Early | N | Y | Y | SRR41050755, SRR41050744 |
| SCHI0093 | T3N2M1c | LUSC | 40 | 71-75 | F | IVB | Late | Y | Y | Y | SRR41050842, SRR41050686, SRR41050675 |
| SCHI0094 | T2aN2M1b | LUSC | 30 | 71-75 | M | IVB | Late | Y | Y | Y | SRR41050943, SRR41050932, SRR41050738 |
| SCHI0097 | T2aN3M0 | LUSC | 30 | 86-90 | M | IIIC | Late | N | Y | Y | SRR41050727, SRR41050725 |
| SCHI0104 | T3N1M0 | LUSC | 60 | 81-85 | M | IIIA | Late | N | Y | Y | SRR41050724, SRR41050723 |
| SCHI0106 | T4N3M1c | LUAD | 50 | 61-65 | F | IVB | Late | N | Y | Y | SRR41050719, SRR41050718 |
| SCHI0110 | T2aN3M0 | NSCLC | 50 | 71-75 | M | IIIB | Late | Y | N | Y | SRR41050717, SRR41050716, SRR41050835 |
| SCHI0113 | T3N0M0 | LUAD | 50 | 66-70 | M | IIB | Early | Y | Y | Y | SRR41050834, SRR41050833, SRR41050832 |
| SCHI0120 | T2bN0M0 | LUAD | 30 | 81-85 | M | IIA | Early | Y | Y | Y | SRR41050831, SRR41050829, SRR41050828 |
| SCHI0122 | T2bN0M0 | LUAD | 40 | 71-75 | F | IIA | Early | Y | Y | Y | SRR41050827, SRR41050826, SRR41050825 |
| SCHI0137 | T2bN0M0 | LUAD | 20 | 71-75 | M | IIA | Early | Y | Y | Y | SRR41050824, SRR41050823, SRR41050822 |
| SCHI0138 | T1cN0M0 | LUAD | 10 | 61-65 | F | IA3 | Early | Y | Y | Y | SRR41050821, SRR41050820, SRR41050818 |
| SCHI0139 | T4N3M1c | LUAD | 30 | 61-65 | M | IVB | Late | Y | Y | Y | SRR41050817, SRR41050816, SRR41050815 |
| SCHI0142 | T2bN0M0 | LUSC | 20 | 71-75 | M | IIA | Early | Y | Y | Y | SRR41050814, SRR41050813, SRR41050812 |
| SCHI0144 | T1cN0M0 | LUAD | 10 | 66-70 | F | IA3 | Early | Y | Y | Y | SRR41050667, SRR41050666, SRR41050665 |
| SCHI0145 | T4N2M1a | LUSC | 20 | 66-70 | M | IVA | Late | Y | Y | Y | SRR41050663, SRR41050662, SRR41050661 |
| SCHI0147 | T2aN0M0 | LUSC | 15 | 81-85 | M | IB | Early | Y | Y | Y | SRR41050660, SRR41050659, SRR41050658 |
| SCHI0155 | T1cN0M0 | LUAD | 20 | 66-70 | F | IA3 | Early | Y | Y | Y | SRR41050657, SRR41050656, SRR41051006 |
| SCHI0156 | T1bN0M0 | LUAD | 50 | 71-75 | F | IA2 | Early | Y | Y | Y | SRR41051005, SRR41051003, SRR41051002 |
| SCHI0167 | T2aN0M0 | LUAD | 20 | 71-75 | F | IB | Early | Y | Y | Y | SRR41051001, SRR41051000, SRR41050999 |
| SCHI0171 | T3N3M0 | LUSC | 30 | 66-70 | F | IIIB | Late | N | Y | Y | SRR41050998, SRR41050997 |
| SCHI0177 | T2aN0M0 | Carcinoid | NA | 76-80 | F | IB | Early | N | Y | Y | --- |
| SCHI0192 | T4N2M0 | LUSC | 50 | 71-75 | M | IIIB | Late | Y | Y | Y | SRR41050996, SRR41050995, SRR41050922 |
| SCHI0193 | T2N2M0 | LUSC | 40 | 71-75 | F | IIIB | Late | Y | Y | Y | SRR41050920, SRR41050919, SRR41050918 |
| SCHI0194 | T4N0M0 | LUSC | 55 | 66-70 | M | IIIA | Late | Y | Y | Y | SRR41050917, SRR41050916, SRR41050915 |
| SCHI0195 | T3N0M0 | LUSC | 25 | 61-65 | M | IIB | Early | Y | Y | Y | SRR41050914, SRR41050913, SRR41050912 |
| SCHI0197 | T2aN3M0 | LUAD | 45 | 61-65 | F | IIIB | Late | Y | Y | Y | SRR41050911, SRR41050811, SRR41050809 |
| SCHI0198 | T4N2M0 | SCLC | 40 | 61-65 | M | IIIB | Late | Y | Y | Y | SRR41050808, SRR41050807, SRR41050806 |
| SCHI0199 | T1bN0M0 | LUAD | 15 | 66-70 | M | IA2 | Early | Y | Y | Y | SRR41050805, SRR41050804, SRR41050803 |
| SCHI0200 | T1cN2M1b | SCLC | 50 | 76-80 | F | IVA | Late | Y | Y | Y | SRR41050802, SRR41050800, SRR41050799 |
| SCHI0201 | T2aN0M0 | LUSC | 60 | 76-80 | F | IB | Early | Y | Y | Y | SRR41050798, SRR41050797, SRR41050796 |
| SCHI0202 | T3N2M0 | SCLC | 10 | 76-80 | F | IIIB | Late | Y | Y | Y | SRR41050795, SRR41050794, SRR41050792 |
| SCHI0203 | T1cN0M0 | LUSC | 50 | 66-70 | M | IA3 | Early | Y | Y | Y | SRR41050791, SRR41050789, SRR41050788 |
| SCHI0204 | T3N2M1c | NSCLC | 15 | 76-80 | F | IVB | Late | Y | Y | Y | SRR41050907, SRR41050906, SRR41050905 |
| SCHI0205 | T1cN2M1c | SCLC | 50 | 61-65 | F | IVB | Late | Y | Y | Y | SRR41050904, SRR41050903, SRR41050901 |
| SCHI0206 | T2bN3M0 | LUAD | 50 | 66-70 | M | IIIB | Late | Y | Y | Y | SRR41050900, SRR41050898, SRR41050897 |
| SCHI0207 | T4N2M1c | SCLC | 70 | 76-80 | M | IVB | Late | Y | Y | Y | SRR41050896, SRR41050895, SRR41050894 |
| SCHI0208 | T1cN1M0 | NSCLC | 40 | 66-70 | F | IIB | Early | Y | Y | Y | SRR41050893, SRR41050892, SRR41050891 |
| SCHI0209 | T4N3M1c | LUAD | NA | 71-75 | M | IVB | Late | Y | Y | Y | SRR41050890, SRR41050889, SRR41050886 |
| SCHI0210 | T2aN0M0 | NSCLC | 10 | 76-80 | F | IB | Early | Y | Y | Y | SRR41050885, SRR41050884, SRR41050714 |
| SCHI0211 | T2bN2M1a | LUAD | 20 | 71-75 | F | IVA | Late | Y | Y | Y | SRR41050713, SRR41050712, SRR41050710 |
| SCHI0212 | T3N1M1c | NSCLC | 20 | 71-75 | M | IVB | Late | Y | Y | Y | SRR41050708, SRR41050707, SRR41050705 |
| SCHI0213 | T1bN0M0 | NSCLC | 20 | 56-60 | M | IA2 | Early | Y | Y | Y | SRR41050704, SRR41050703, SRR41050702 |
| SCHI0214 | T1cN0M0 | NSCLC | 50 | 71-75 | F | IA3 | Early | Y | Y | Y | SRR41050701, SRR41050700, SRR41050699 |
| SCHI0218 | T4N31b | SCLC | 20 | 76-80 | F | IVA | Late | Y | Y | Y | SRR41050697, SRR41050696, SRR41050695 |
| SCHI0219 | T1bN1M0 | NSCLC | 30 | 66-70 | M | IIb | Early | Y | Y | Y | SRR41050694, SRR41050693, SRR41050692 |
| SCHI0220 | T3N3M1b | LUAD | 40 | 71-75 | F | IVA | Late | Y | Y | Y | SRR41050994, SRR41050993, SRR41050992 |
| SCHI0222 | T4N1M0 | LUAD | 55 | 66-70 | M | IIIA | Late | Y | Y | Y | SRR41050991, SRR41050989, SRR41050988 |
| SCHI0223 | T4N3M1c | SCLC | 50 | 71-75 | M | IVB | Late | Y | Y | Y | SRR41050987, SRR41050986, SRR41050985 |
| SCHI0224 | T2bN2M0 | LUSC | 60 | 61-65 | M | IIIA | Late | Y | Y | Y | SRR41050984, SRR41050983, SRR41050981 |
| SCHI0227 | T2bN0M0 | NSCLC | 50 | 86-90 | M | IIB | Early | Y | Y | Y | SRR41050980, SRR41050978, SRR41050977 |
| SCHI0228 | T1bN3M1c | LUAD | 20 | 71-75 | M | IVB | Late | Y | Y | Y | SRR41050976, SRR41050975, SRR41050974 |
| SCHI0229 | T4N0M1a | NSCLC | NA | 86-90 | M | IVA | Late | Y | Y | Y | SRR41050973, SRR41050972, SRR41050971 |
| SCHI0232 | T2aN2M0 | NSCLC | 20 | 71-75 | M | IIIA | Late | Y | Y | Y | SRR41050787, SRR41050786, SRR41050784 |
| SCHI0233 | T4N2M0 | SCLC | 40 | 61-65 | F | IIIA | Late | Y | Y | Y | SRR41050783, SRR41050782, SRR41050781 |
| SCHI0234 | T1cN0M0 | LUSC | 60 | 76-80 | M | IA3 | Early | Y | Y | Y | SRR41050780, SRR41050779, SRR41050778 |
| SCHI0238 | T2aN0M0 | LUAD | 51 | 61-65 | F | IB | Early | Y | Y | Y | SRR41050776, SRR41050775, SRR41050772 |
| SCHI0241 | T2aN31b | LUAD | 25 | 81-85 | M | IVA | Late | Y | Y | Y | SRR41050771, SRR41050770, SRR41050769 |
| SCHI0242 | T3N0M0 | LUSC | 40 | 71-75 | M | IIB | Early | Y | Y | Y | SRR41050768, SRR41050767, SRR41050766 |
| SCHI0244 | T3N0M1c | LUSC | 20 | 51-55 | F | IVB | Late | Y | Y | Y | SRR41050765, SRR41050764, SRR41050881 |
| SCHI0245 | T2bN0M0 | LUSC | 30 | 61-65 | M | IIA | Early | Y | Y | Y | SRR41050880, SRR41050879, SRR41050877 |
| SCHI0249 | T2aN2M0 | LUAD | 30 | 66-70 | M | IIIA | Late | Y | Y | Y | SRR41050875, SRR41050874, SRR41050872 |
| SCHI0251 | T4N3M1c | NSCLC | 40 | 61-65 | M | IVB | Late | Y | Y | Y | SRR41050870, SRR41050869, SRR41050867 |
| SCHI0252 | T1cN2M0 | NSCLC | 20 | 66-70 | F | IIIA | Late | Y | Y | Y | SRR41050866, SRR41050865, SRR41050863 |
| SCHI0253 | T1cN3M1b | NSCLC | 15 | 76-80 | F | IVA | Late | Y | Y | Y | SRR41050862, SRR41050861, SRR41050969 |
| SCHI0259 | T3N2M0 | LUAD | 40 | 71-75 | M | IIIB | Late | Y | Y | Y | SRR41050968, SRR41050967, SRR41050965 |
| SCHI0265 | T2bN0M0 | NSCLC | 33 | 61-65 | M | IIA | Early | Y | Y | Y | SRR41050964, SRR41050963, SRR41050962 |
| SCHI0266 | T2aN0M0 | NSCLC | 30 | 71-75 | F | IB | Early | Y | Y | Y | SRR41050961, SRR41050959, SRR41050958 |
| SCHI0272 | T1cN0M0 | NSCLC | 40 | 76-80 | F | IA3 | Early | Y | Y | Y | SRR41050955, SRR41050954, SRR41050953 |
| SCHI0275 | T2aN0M0 | LUSC | 20 | 76-80 | F | IB | Early | Y | Y | Y | SRR41050952, SRR41050951, SRR41050950 |
| SCHI0276 | T1cN1M0 | SCLC | 30 | 66-70 | M | IIB | Early | Y | Y | Y | SRR41050948, SRR41050947, SRR41050762 |
| SCHI0277 | T1cN3M1b | LUAD | 20 | 76-80 | F | IVA | Late | Y | Y | Y | SRR41050761, SRR41050760, SRR41050758 |
| SCHI0280 | T1c2bN1c | LUAD | NA | 61-65 | F | IVB | Late | Y | Y | Y | SRR41050757 |
| SCHI0281 | T3N1M0 | LUAD | 15 | 66-70 | M | IIIA | Late | Y | Y | Y | SRR41050756, SRR41050754, SRR41050753 |
| SCHI0282 | T3N0M0 | NSCLC | 30 | 61-65 | F | IIB | Early | Y | Y | Y | SRR41050752, SRR41050751, SRR41050750 |
| SCHI0283 | T2aN0M0 | NSCLC | 30 | 81-85 | M | IB | Early | Y | Y | Y | SRR41050749, SRR41050748, SRR41050746 |
| SCHI0285 | T1bN0M0 | LUAD | 40 | 66-70 | M | IA | Early | Y | Y | Y | SRR41050745, SRR41050743, SRR41050741 |
| SCHI0286 | T3N1M0 | LUAD | 40 | 71-75 | F | IIIA | Late | Y | Y | Y | SRR41050740, SRR41050859, SRR41050858 |
| SCHI0288 | T2aN0M0 | LUAD | 40 | 56-60 | F | IB | Early | Y | Y | Y | SRR41050857, SRR41050856, SRR41050855 |
| SCHI0289 | T4N3M0 | LUAD | 50 | 66-70 | F | IIIC | Late | Y | Y | Y | SRR41050854, SRR41050851 |
| SCHI0290 | T2bN0M0 | LUSC | 40 | 71-75 | M | IIA | Early | Y | Y | Y | SRR41050850, SRR41050849, SRR41050848 |
| SCHI0291 | T2aN0M0 | LUAD | 35 | 71-75 | M | IB | Early | Y | Y | Y | SRR41050847, SRR41050846, SRR41050844 |
| SCHI0293 | T2N0M0 | LUAD | 15 | 81-85 | F | IIIA | Late | Y | Y | Y | SRR41050841, SRR41050840, SRR41050838 |
| SCHI0294 | T1cN0M0 | LUAD | 20 | 71-75 | F | IA3 | Early | Y | Y | Y | SRR41050837, SRR41050836, SRR41050691 |
| SCHI0295 | T1bN2aM0 | LUAD | NA | 66-70 | F | IIA | Early | Y | Y | Y | SRR41050690, SRR41050689, SRR41050688 |
| SCHI0300 | T1c2bM0 | SCLC | 40 | 66-70 | F | IIIA | Late | Y | Y | Y | SRR41050683, SRR41050682, SRR41050681 |
| SCHI0303 | T2aN0M0 | SCLC | 30 | 51-55 | F | IB | Early | Y | Y | Y | SRR41050680, SRR41050679, SRR41050678 |
| SCHI0305 | T4N2M1C | NSCLC | 15 | 76-80 | M | IVB | Late | Y | Y | Y | SRR41050677, SRR41050676 |
| SCHI0314 | T2qN0M0 | LUSC | 10 | 81-85 | F | IB | Early | Y | Y | Y | SRR41050674, SRR41050673, SRR41050672 |
| SCHI0315 | T1cN3M1b | LUAD | 40 | 61-65 | M | IVA | Late | Y | Y | Y | SRR41050671, SRR41050670, SRR41050669 |
| SCHI0316 | T2bN0M0 | LUAD | 5 | 56-60 | F | IIA | Early | Y | Y | Y | SRR41050668, SRR41050946, SRR41050945 |
| SCHI0319 | T1bN0M0 | NSCLC | 20 | 66-70 | M | IA2 | Early | Y | Y | Y | SRR41050942, SRR41050941, SRR41050940 |
| SCHI0342 | T2bN0M0 | LUAD | 10 | 66-70 | F | IIA | Early | Y | Y | Y | SRR41050933, SRR41050931 |
| SCHI0343 | T1bN0M0 | LUAD | 50 | 66-70 | M | IA2 | Early | Y | Y | Y | SRR41050930, SRR41050929 |
| SCHI0344 | T1cN0M0 | LUSC | 60 | 66-70 | F | IA3 | Early | Y | Y | Y | SRR41050928 |
| SCHI0346 | T4N0M0 | NSCLC | 30 | 51-55 | M | IIIA | Late | Y | Y | Y | SRR41050927, SRR41050926 |
| SCHI0350 | T4N0M0 | LUSC | 30 | 76-80 | M | IIIA | Late | Y | Y | Y | SRR41050923, SRR41050739, SRR41050736 |
| SCHI0351 | T2bN0M0 | NSCLC | 50 | 81-85 | M | IIA | Early | Y | Y | Y | SRR41050735, SRR41050734, SRR41050733 |

Abbreviations: BW. Bronchial washing; LUAD, lung adenocarcinoma; LUSC, lung squamous cell carcinoma; N, no (not collected); NA, not applicable; SCLC, small-cell lung cancer; TBLB, transbronchial forceps biopsy of lung tumour tissue; EBUS-TBNA, endobronchial ultrasound-guided transbronchial needle aspirate of the lung tumour tissue; transbronchial needle aspiration; TNM (system for lung cancer staging), Tumour, Node, and Metastasis; Y, yes (collected). Green highlighting: specimen subjected to metagenomic sequencing; Blue highlighting: specimen excluded from metagenomic sequencing due to insufficient microbial DNA; Purple highlighting; specimen yielded no detectable microbiome upon metagenomic sequencing; Light grey highlighting: no specimen collected, or (for scopes) surplus specimens were collected. *Chronic obstructive pulmonary disease co-morbidity. ^†^Human immunodeficiency virus-1 co-morbidity.

**Table S2. Healthy control participants, collected specimens, and metagenome-sequenced specimens.**

| **Participant** | **Age Range** | **Sex** | **Smoking Pack-Years** | **Oral Wash** | **BW** | **SRA IDs** |
| --- | --- | --- | --- | --- | --- | --- |
| ***Healthy controls providing bronchial washes (BWs)*** | | | | | | |
| SCHI0105 | 81-85 | M | 10 | N | Y* | SRR41050722, SRR41050720 |
| SCHI0267 | 66-70 | F | 5 | Y | Y | SRR41050957, SRR41050956 |
| SCHI0318 | 81-85 | M | 10 | Y | Y | SRR41050944 |
| SCHI0326 | 21-25 | F | NA | Y | Y | SRR41050939, SRR41050938 |
| SCHI0327 | 61-65 | M | 10 | Y | Y | SRR41050937, SRR41050936 |
| SCHI0328 | 51-55 | F | NA | Y | Y | SRR41050935, SRR41050934 |
| SCHI0347 | 81-85 | F | 5 | Y | Y | SRR41050925, SRR41050924 |
| SCHI0352 | 71-75 | M | 10 | Y | Y | SRR41050732, SRR41050731 |
| SCHI0353 | 61-65 | F | NA | Y | Y | SRR41050730, SRR41050729 |
| SCHI0358 | 81-85 | M | NA | Y | Y | SRR41050728, SRR41050726 |
| ***Healthy controls providing oral washes only*** | | | | | | |
| SCHI0026 | 41-45 | M | 2 | Y | N | SRR41050921 |
| SCHI0092 | 41-45 | F | 0 | Y | N | SRR41050853 |
| SCHI0236 | 26-30 | F | 0 | Y | N | SRR41050777 |
| SCHI0248 | 41-45 | M | 0 | Y | N | SRR41050876 |
| SCHI0292 | 26-30 | F | 0 | Y | N | SRR41050843 |
| SCHI0296 | 21-25 | F | 0 | Y | N | SRR41050685 |

*Due to initial suspicion of lung cancer in this participant, two bronchial washes from each lung were obtained.

Green highlighting: specimen subjected to metagenomic sequencing; Blue highlighting: specimen excluded from metagenomic sequencing due to insufficient microbial DNA; light grey highlighting: no specimen collected, or (for scopes) surplus specimens were collected.

**Table S3. Reagent-only and bronchoscope wash metagenome-sequenced samples.**

| **Study ID** | **Control type** | **SRA ID** |
| --- | --- | --- |
| SCHI0197_Scope_Stage_IIIB_LUAD | Bronchoscope wash | SRR41050810 |
| SCHI0202_Scope_Stage_IIIB_SCLC | Bronchoscope wash | SRR41050793 |
| SCHI0205_Scope_Stage_IVB_SCLC | Bronchoscope wash | SRR41050902 |
| SCHI0209_Scope_Stage_IVB_LUAD | Bronchoscope wash | SRR41050887 |
| SCHI0210_Scope_Stage_IB_NSCLC-NOS | Bronchoscope wash | SRR41050715 |
| SCHI0211_Scope_Stage_IVA_LUAD | Bronchoscope wash | SRR41050711 |
| SCHI0212_Scope_Stage_IVB_NSCLC-NOS | Bronchoscope wash | SRR41050706 |
| SCHI0224_Scope_Stage_IIIA_LUSC | Bronchoscope wash | SRR41050982 |
| SCHI0244_Scope_Stage_IVB_LUSC | Bronchoscope wash | SRR41050883 |
| SCHI0245_Scope_Stage_IIA_LUSC | Bronchoscope wash | SRR41050878 |
| SCHI0249_Scope_Stage_IIIA_LUAD | Bronchoscope wash | SRR41050873 |
| SCHI0251_Scope_Stage_IVB_NSCLC-NOS | Bronchoscope wash | SRR41050868 |
| SCHI0252_Scope_Stage_IIIA_NSCLC-NOS | Bronchoscope wash | SRR41050864 |
| SCHI0253_Scope_Stage_IVA_NSCLC-NOS | Bronchoscope wash | SRR41050970 |
| SCHI0259_Scope_Stage_IIIB_LUAD | Bronchoscope wash | SRR41050966 |
| SCHI0276_Scope_Stage_IIB_SCLC | Bronchoscope wash | SRR41050763 |
| SCHI0277_Scope_Stage_IVA_LUAD | Bronchoscope wash | SRR41050759 |
| SCHI0283_Scope_Stage_IB_NSCLC-NOS | Bronchoscope wash | SRR41050747 |
| SCHI0285_Scope_Stage_IA_LUAD | Bronchoscope wash | SRR41050742 |
| SCHI0289_Scope_Stage_IIIC_LUAD | Bronchoscope wash | SRR41050852 |
| SCHI0291_Scope_Stage_IB_LUAD | Bronchoscope wash | SRR41050845 |
| SCHI0293_Scope_Stage_IIIA_LUAD | Bronchoscope wash | SRR41050839 |
| SCHI0297_Scope_Lymphoma | Bronchoscope wash | SRR41050684 |
| SCHI0350_Scope_Stage_IIIA_LUSC | Bronchoscope wash | SRR41050737 |
| NTC_NEG_CONTROL1 | Reagent-only control | SRR41050910 |
| NTC_NEG_CONTROL2 | Reagent-only control | SRR41050909 |
| NTC_NEG_CONTROL3 | Reagent-only control | SRR41050774 |
| NTC_NEG_CONTROL4 | Reagent-only control | SRR41050687 |
| NTC_NEG_CONTROL5 | Reagent-only control | SRR41050721 |
| NTC_NEG_CONTROL6 | Reagent-only control | SRR41050830 |
| NTC_NEG_CONTROL7 | Reagent-only control | SRR41050819 |
| NTC_NEG_CONTROL8 | Reagent-only control | SRR41050664 |
| NTC_NEG_CONTROL9 | Reagent-only control | SRR41051004 |

**Supplementary Figures**

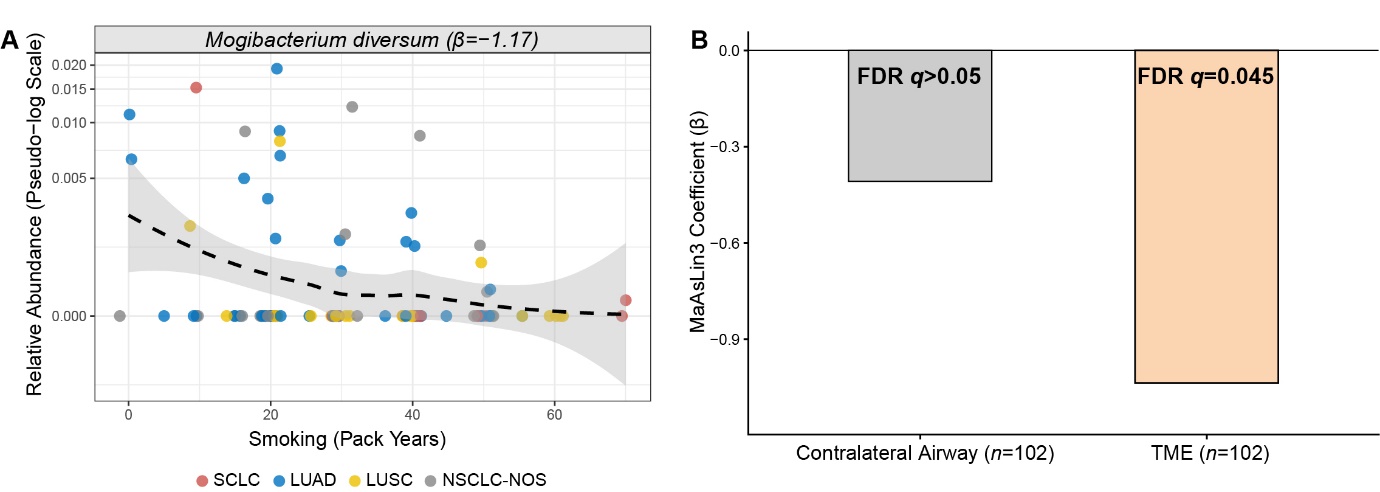

**Figure S1. Impact of cumulative tobacco smoking on the microbiome of the lung cancer tumour microenvironment (TME).** (A) Cross-sectional Maaslin3 multivariable linear modelling of TME microbiota identified significant, progressive depletion of the commensal bacterium, *Mogibacterium diversum* (β coefficient=-1.17; false discovery rate [FDR]-adjusted *q*=0.048) in response to increasing smoking pack-years. This microbial biomarker remained significant after controlling for lung cancer subtypes (small-cell lung carcinoma [SCLC], lung adenocarcinoma [LUAD], lung squamous cell carcinoma [LUSC], and non-small cell lung carcinoma not otherwise specified [NSCLC-NOS]). (B) Fixed-effects modelling revealed that *M. diversum* depletion was highly pronounced within the TME (β coefficient=-1.17; *q*=0.048), but not the matching contralateral airway (β coefficient=-0.40; *q*=0.98). This finding indicates that *M. diversum* depletion is associated with both cumulative smoking pack-years and tumour presence, rather than being a generalised smoking or tumour signature alone.

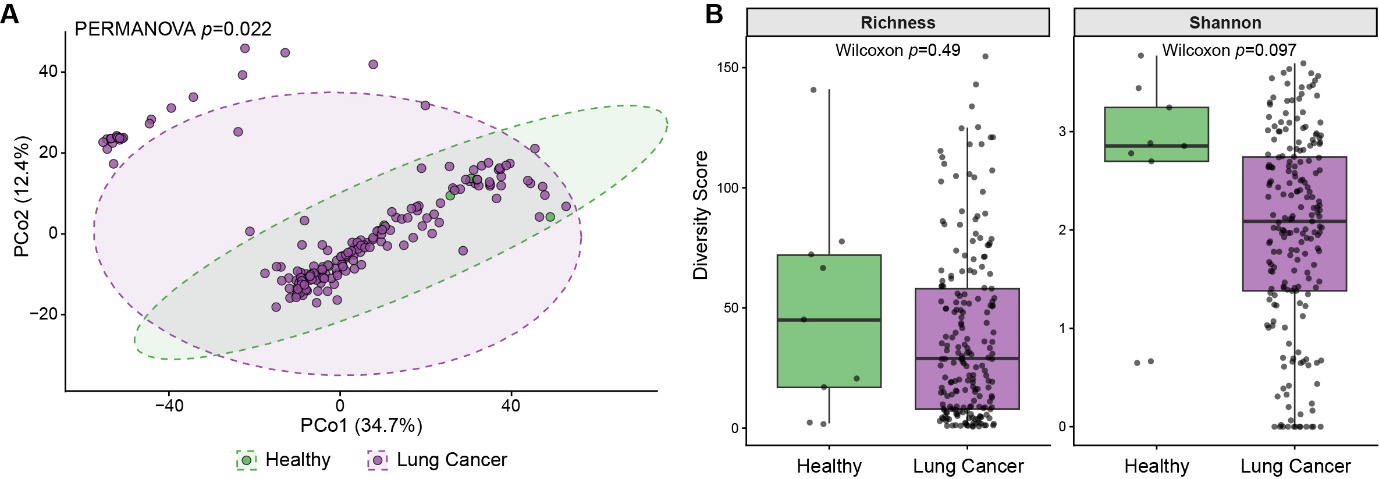

**Figure S2. Global disruption of the lung microbiome in participants with lung cancer.** (A) Principal Coordinate Analysis was used to visualise lung microbiome β diversity differences between healthy controls and the combined lung cancer cohort (i.e. both tumour and paired contralateral bronchial washes [BWs]). Distances were calculated using Aitchison distance (Euclidean distance of centred log-ratio transformed abundances). Ellipses denote the 95% confidence intervals for each clinical group. Multivariable PERMANOVA, adjusted for participant smoking history (pack-years), demonstrated a significant global divergence in lung cancer-driven microbial community architecture (PERMANOVA *p*=0.022; beta-dispersion *p*=0.678). This non-significant beta-dispersion confirms that the global divergence represents a genuine compositional shift between healthy and diseased airways, rather than unequal variance. (B) Boxplots displaying Richness and Shannon diversity index, which identified no significant α diversity differences between the lung cancer and healthy cohorts (*p*>0.05). Significance was assessed using an unpaired two-sample Wilcoxon rank-sum test. Solid black lines represent the median, and boxes span the interquartile range.

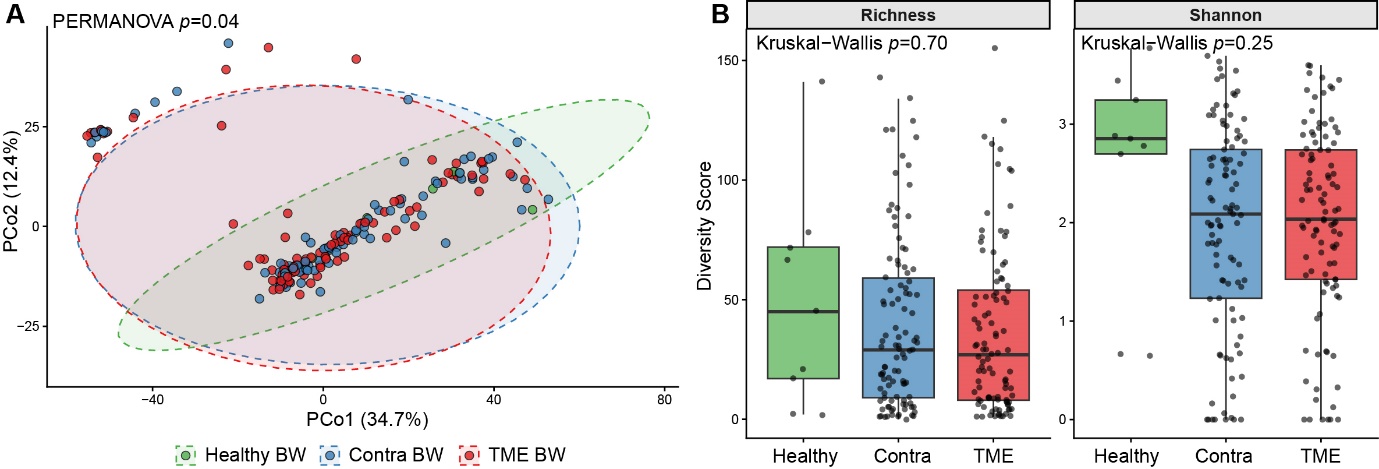

**Figure S3. Impact of anatomical site on the lung microbiome in participants with lung cancer.** (A) Principal Coordinate Analysis was used to visualise lung microbiome β diversity differences between healthy controls and paired contralateral (Contra) and tumour microenvironment (TME) BWs from lung cancer participants. Distances were calculated using Aitchison distance (Euclidean distance of centred log-ratio transformed abundances). Ellipses denote the 95% confidence intervals for each clinical group. Multivariable PERMANOVA, adjusted for participant smoking history (pack-years), demonstrated significant overall microbial community structural divergence (PERMANOVA *p*=0.04; beta-dispersion *p*=0.743). The non-significant beta-dispersion confirms this global divergence is driven by a genuine compositional shift rather than unequal variance between groups. However, subsequent post-hoc pairwise PERMANOVA with Benjamini-Hochberg false discovery rate correction failed to identify distinct microbial community architectures between anatomical sites (Healthy vs. Contra *p*=0.068; Healthy vs. TME *p*=0.092; Contra vs. TME *p*=0.068). (B) Boxplots displaying Richness and Shannon diversity metrics, comparing Healthy BWs to paired Contra and TME BWs. Global ecological stability was assessed using the Kruskal-Wallis omnibus test, which identified no significant differences in overall α diversity (*p*>0.05). The solid black lines represent the median, with boxes spanning the interquartile range.

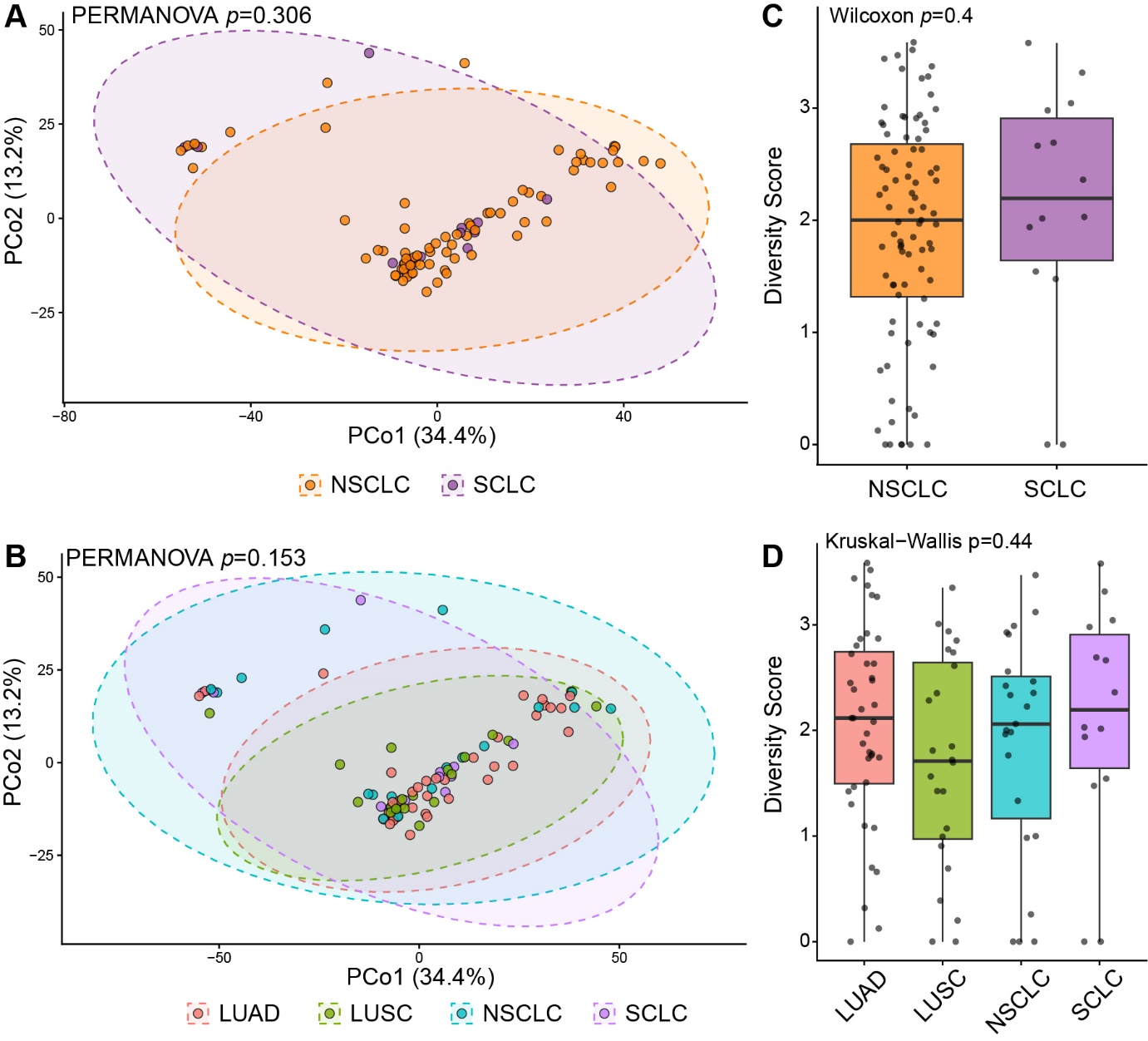

**Figure S4. Impact of lung cancer subtype on the lung microbiome in participants with lung cancer.** (A) Principal Coordinate Analysis was used to visualise lung microbiome β diversity differences between non-small cell lung carcinoma (NSCLC) and small cell lung cancer (SCLC) from lung cancer participants. Distances were calculated using Aitchison distance (Euclidean distance of centred log-ratio transformed abundances). Ellipses denote the 95% confidence intervals for each clinical group. Multivariable PERMANOVA, adjusted for participant smoking history (pack-years), demonstrated no microbial community structural divergence (PERMANOVA *p*>0.05; beta-dispersion *p>*0.05). (B) Same visualisation as (A), but with more granular NSCLC subtype assessment for lung adenocarcinoma (LUAD), lung squamous cell carcinoma (LUSC), and non-small cell lung carcinoma not otherwise specific (NSCLC-NOS). No microbial community structural divergence was seen (PERMANOVA *p*>0.05; beta-dispersion *p*>0.05). (C) and (D) Boxplots displaying the Shannon diversity index, comparing cohorts from (A) and (B), respectively. Both analyses identified no significant differences in α diversity (*p*>0.05). The solid black lines represent the median, with boxes spanning the interquartile range.

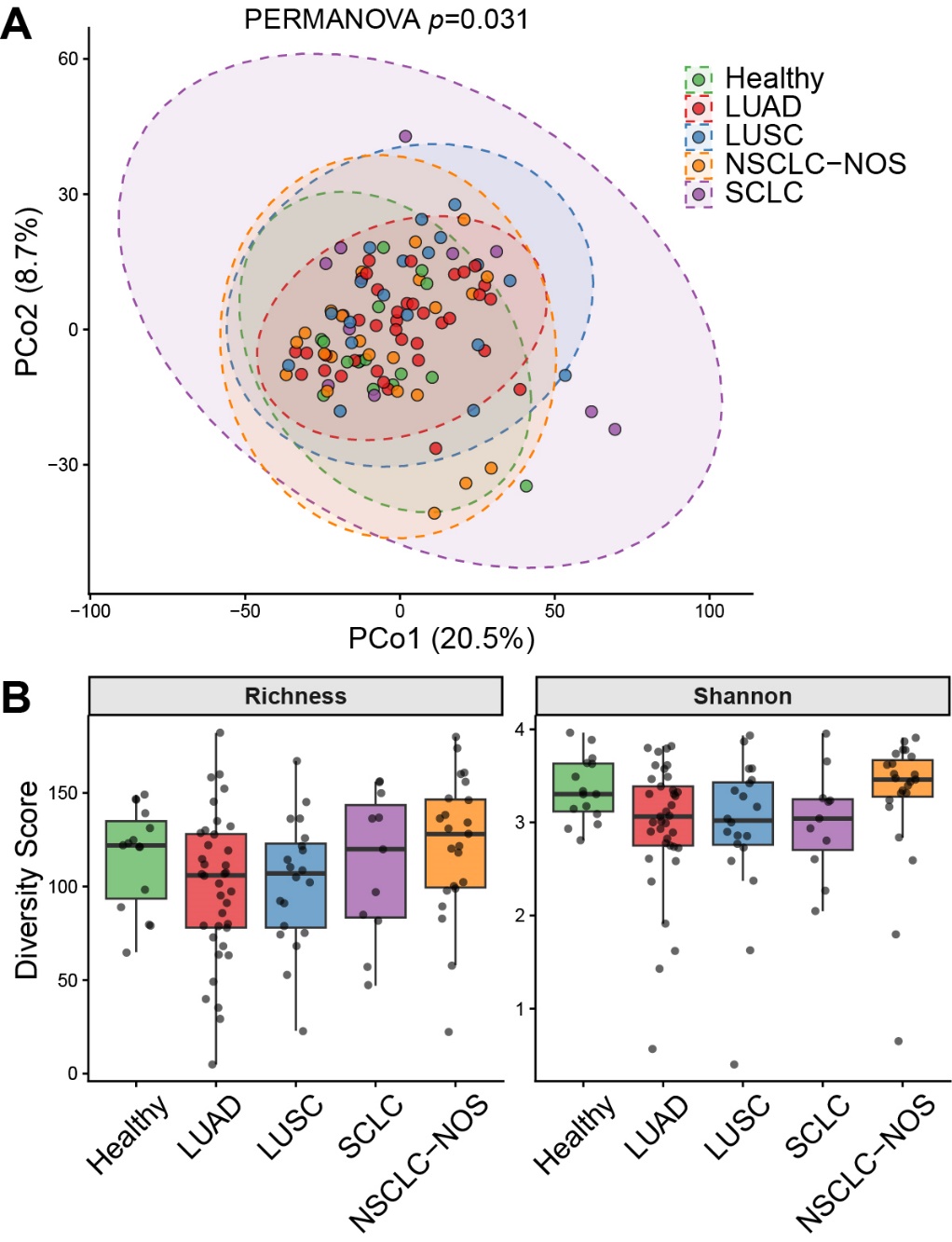

**Figure S5. Oral microbiome community structure is significantly altered in lung adenocarcinoma (LUAD), but not other histological subtypes.** (A) Principal Coordinate Analysis based on Aitchison distance (Euclidean distance of centred log-ratio transformed abundances) was used to visualise oral microbiome β diversity across healthy controls and lung cancer subtypes. Ellipses represent 95% confidence intervals for each cohort. Multivariable PERMANOVA, adjusted for participant smoking history (pack-years), identified a significant global oral β diversity difference (PERMANOVA *p*=0.031; beta-dispersion *p*=0.046). The concurrent significant dispersion indicates this overarching divergence is driven, at least in part, by unequal ecological destabilisation across the different cancer subtypes compared to the Healthy baseline. Subsequent post-hoc pairwise PERMANOVA with Benjamini-Hochberg false discovery rate (FDR) correction revealed that these structural shifts were primarily driven by the LUAD cohort, which diverged significantly from both Healthy controls (FDR *p*=0.020) and lung squamous cell carcinoma (LUSC; FDR *p*=0.050). In contrast, no significant pairwise β diversity differences were observed for the small cell lung carcinoma (SCLC) or non-small cell lung carcinoma-not otherwise specified (NSCLC-NOS) cohorts. (B) Boxplots displaying oral microbiome Richness and Shannon α diversity. Kruskal-Wallis omnibus testing identified no significant ecological differences across cohorts (all *p*>0.05). Solid black lines represent the median, with boxes spanning the interquartile range.

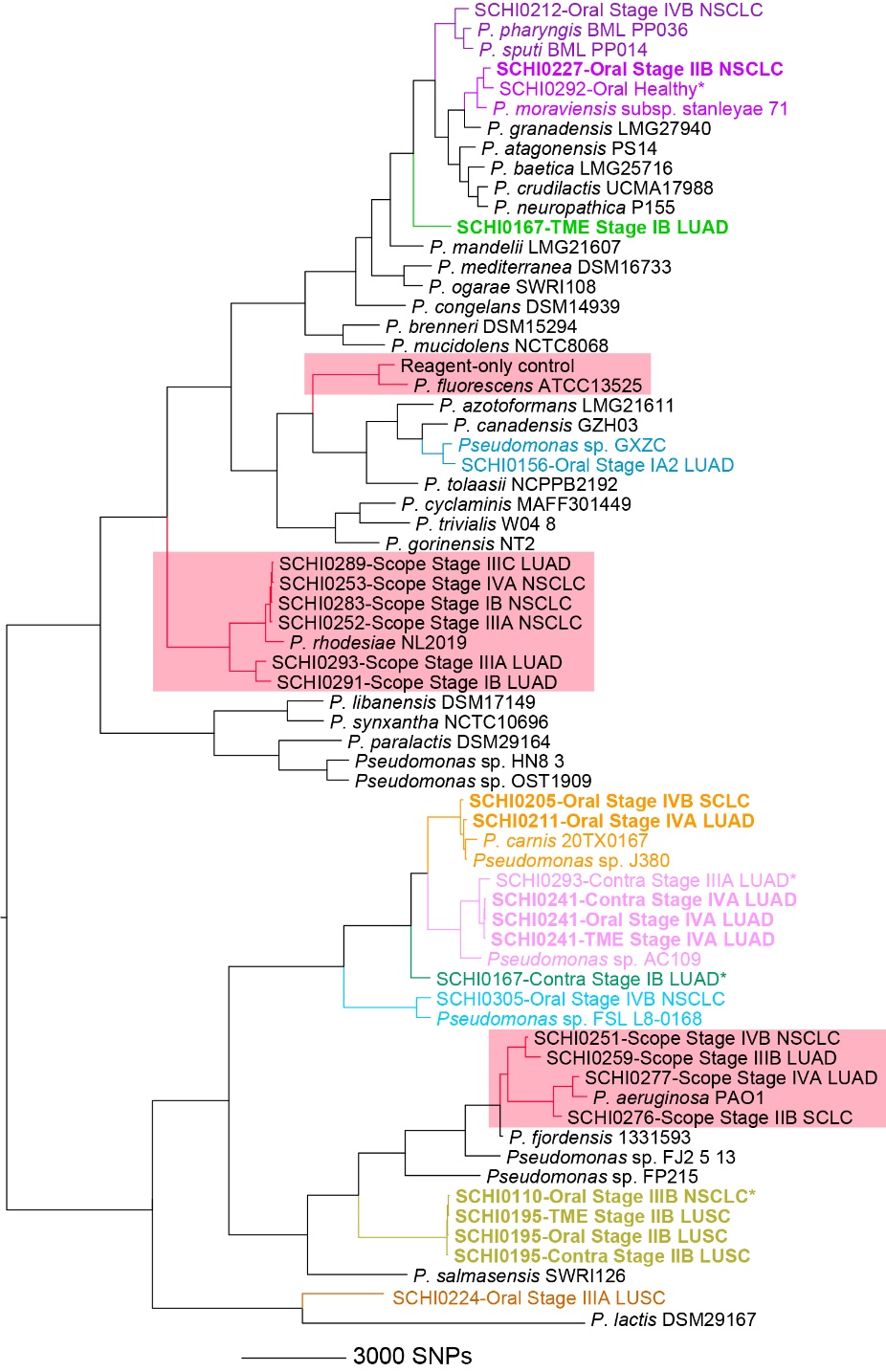

**Figure S6. Midpoint-rooted maximum parsimony phylogenomic reconstruction of *Pseudomonas fluorescens* group (PFG) species, and *Pseudomonas aeruginosa*, detected by Kraken2 in Australian lung cancer participant metagenomes**. To determine whether the enriched *Pseudomonas trivialis* signal observed in tumour microenvironment (TME) bronchial washings (BWs) is supported by comparative genomics, we compared PFG*-*positive TME BWs, contralateral (Contra) BWs, and oral washes with high-abundance signal (>29,000 Kraken2-mapped *Pseudomonas* spp. reads) against the *P. lactis* DSM 29167 reference genome using SPANDx v4.3.1. Negative controls (reagent-only controls or bronchoscope washes) meeting this *Pseudomonas* spp. read threshold were also included (highlighted red boxes). This analysis uncovered several PFG taxa in our dataset, including probable known species belonging to previously characterised *Pseudomonas* spp. (purple, blue, orange, pink, and gold clades), along with several potentially novel pseudomonad species (green, gold, and brown clades). This analysis confirms that there is greater pseudomonad diversity present in lung cancer participant-derived specimens than is being accurately resolved by the Kraken2 Standard database. *Denotes *P. trivialis*-negative metagenomes according to Kraken2. Bolded text denotes very high-level *Pseudomonas* signal (>1 million reads) in these specimens, of which a small fraction (~0.1%) were assigned to *P. trivialis* by Kraken2. The scale bar indicates the number of single-nucleotide polymorphisms (SNPs). Abbreviations: LUAD, lung adenocarcinoma; LUSC, lung squamous cell carcinoma; SCLC, small cell lung carcinoma. Consistency index=0.61.
